# Cost-Effectiveness of Interventions to Improve HIV Pre-Exposure Prophylaxis Initiation, Adherence, and Persistence among Men Who Have Sex with Men

**DOI:** 10.1101/2021.07.22.21260930

**Authors:** Margo M. Wheatley, Gregory Knowlton, Szu-Yu Kao, Samuel M. Jenness, Eva Enns

**Affiliations:** Division of Health Policy and Management, University of Minnesota, Minneapolis, MN; Department of Epidemiology, Rollins School of Public Health, Emory University, Atlanta, GA

**Keywords:** cost-effectiveness, pre-exposure prophylaxis, HIV/AIDS

## Abstract

**Background:** To help achieve *Ending the HIV Epidemic* (EHE) goals of reducing new HIV incidence, pre-exposure prophylaxis (PrEP) use and engagement must increase despite multidimensional barriers to scale-up and limitations in funding. We investigated the cost-effectiveness of interventions spanning the PrEP continuum of care for men who have sex with men (MSM) in Atlanta, Georgia, a focal jurisdiction for the *EHE* plan.

**Methods:** Using a network-based HIV transmission model, we simulated lifetime costs, quality-adjusted life years (QALYs), and infections averted for eight intervention strategies using a health sector perspective. Strategies included a status quo (no interventions), three distinct interventions (targeting PrEP initiation, adherence, or persistence), and all possible intervention combinations. Cost-effectiveness was evaluated incrementally using a $100,000/QALY gained threshold. We performed sensitivity analyses on PrEP costs, intervention costs, and intervention coverage.

**Findings:** Strategies averted 0.2–4.2% new infections and gained 0.0045%–0.24% QALYs compared to the status quo. Initiation strategies achieved 20%–23% PrEP coverage (up from 15% with no interventions) and moderate clinical benefits at a high cost, while adherence strategies were relatively low cost and low benefit. Under our assumptions, the adherence and initiation combination strategy was cost-effective with an incremental cost-effectiveness ratio of $86,927/QALY gained. Sensitivity analyses showed no strategies were cost-effective when intervention costs increased by 60% and the strategy combining all three interventions was cost-effective when PrEP costs decreased to $1,000/month.

**Interpretation:** Under reasonable assumptions of intervention uptake and cost, PrEP initiation interventions achieved moderate public health gains and could be cost-effective. However, these analyses demonstrate that substantial financial resources will be needed to improve the PrEP care continuum towards meeting EHE goals.

**Funding:** US National Institutes of Health

**RESEARCH IN CONTEXT:** *Evidence before this study:* We searched PubMed for articles published between 2010-2020 using the term “((“Costs and Cost Analysis”[Mesh]) OR “Cost-Benefit Analysis”[Mesh] OR cost-effective* OR cost-utility OR “economic evaluation” OR “economic impact”) AND (“HIV”[Mesh] OR HIV OR HIV/AIDS) AND (pre-exposure OR PrEP) AND (MSM OR gay OR bisexual OR GBM),” which yielded 79 results. PrEP (compared to no PrEP) for high-risk MSM is generally found to be cost-effective in the United States and other high-income countries, with some variation in findings. However, evidence on the cost and cost-effectiveness of interventions designed to address barriers to effective PrEP use are lacking. Current studies typically model hypothetical improvements that may not be realistically achieved.

*Added value of this study:* Using a stochastic network-based model of HIV, we projected the potential costs, benefits, and cost-effectiveness of real-world interventions to improve PrEP use through increased initiation, adherence, and persistence among MSM in a US urban center. We found real-world interventions to improve PrEP use could be cost-effective, however they would be expensive and achieve only limited clinical gains.

*Implications of all the available evidence:* Lower PrEP costs would improve the cost-effectiveness of expanding and improving PrEP use. Continued expansion of PrEP coverage beyond current levels will likely involve multiple interventions of increasing intensity and cost to engage harder-to-reach populations. Realistic efforts to end the HIV epidemic may require investments and interventions that are above currently accepted willingness-to-pay thresholds.

## INTRODUCTION

The United States federal HIV plan, *Ending the HIV Epidemic* (EHE), calls for reducing new HIV infections by 90% by 2030.^1^ Pre-exposure prophylaxis (PrEP) is among the strategies recommended for reaching this goal. PrEP is effective at preventing HIV infection and is recommended for individuals at substantial risk of HIV, including men who have sex with men (MSM), who accounted for approximately 69% of new HIV diagnoses in 2018.^2^ PrEP use remains low: the Centers for Disease Control and Prevention (CDC) estimate that 1.2 million people in the US had indications for PrEP in 2018, yet only 18% received PrEP prescriptions.^3^ Barriers to effective PrEP use are complex and occur along a PrEP care continuum that includes access, prescription, initiation, adherence, and persistence or retention.^4^ Gaps in this continuum may help explain why increasing PrEP prescriptions have had limited population-level impact on reducing HIV incidence.^5^

To reach EHE goals, addressing these PrEP continuum gaps is needed. With a fragmented healthcare financing system and limited funding available for improving PrEP coverage and engagement, estimating the expected costs, benefits, and value of PrEP interventions is important for decision makers. Cost-effectiveness analysis provides a standard framework for evaluating the relative value of intervention strategies.^6^ Prior studies have generally shown PrEP to be cost-effective when targeted to high-risk individuals.^7^ Additional studies have projected clinical benefits of hypothetical PrEP expansion, optimal population prioritization, and improved retention.^8–11^ However, the cost-effectiveness of specific, real-world interventions aimed at addressing barriers to PrEP use has not been well-established.

Cost-effectiveness studies of PrEP typically evaluate the cost of expanded PrEP coverage, but not the resource investments needed to realistically achieve PrEP scale-up, such as costs of interventions to improve patient awareness or reduce barriers to access. Ignoring these implementation and scale-up costs may yield overly optimistic cost-effectiveness estimates of expanded PrEP coverage. Furthermore, ignoring the mechanisms by which PrEP coverage expansion occurs may lead to the evaluation of overly ambitious PrEP coverage levels that would be difficult, and potentially very expensive, to achieve in practice, further distorting cost-effectiveness conclusions.

In this study, we explicitly modeled the costs and mechanisms of PrEP expansion using trial-based interventions. Our primary objective was to assess the potential cost-effectiveness of interventions designed to improve initiation, adherence, and persistence on PrEP among MSM, a high-priority population for PrEP, in an EHE target geographic area.

## METHODS

### Study Design

Using a network-based HIV transmission model, we evaluated cost-effectiveness of PrEP care continuum interventions aimed at increasing PrEP initiation, adherence, and persistence from a health sector perspective. We focused on Atlanta, Georgia because it has among the highest rates of HIV diagnoses in US and is one of the geographic focus areas of EHE.^1^ We simulated a population of 12,000 MSM for computational efficiency then extrapolated results to reflect the total population of approximately 103,000 MSM in Atlanta.^12^

### Transmission Model

We used EpiModel, a dynamic transmission model that has previously been described in detail (Appendix, Sections 1-10).^13–15^ Our model was fit to sexual network, HIV care cascade, and PrEP cascade data for MSM in Atlanta. Parameter values were directly estimated from primary data or derived from published literature and calibrated to HIV surveillance data.

Within each weekly time step, individuals who tested negative for HIV and met the CDC behavioral indications for PrEP had a probability of initiating PrEP.^16^ If PrEP was initiated, individuals were assigned a PrEP adherence level (low, medium, high) which impacted the per-act probability of acquiring HIV. Individuals had a weekly probability of spontaneously discontinuing PrEP and would also discontinue if no longer indicated for PrEP, which was reassessed annually. Individuals could restart PrEP if they acquired new indications. If infected with HIV, a PrEP user would discontinue at their first HIV-positive test.

### PrEP Continuum Interventions

We evaluated three interventions targeting PrEP initiation, adherence, and persistence. These interventions supplemented PrEP delivery in a status quo scenario, in which 15% of PrEP-indicated MSM were on PrEP at any given time,^3^ 60% of users had high adherence,^17,18^ and median duration of PrEP use was 227 days.^19,20^

The PrEP initiation intervention was based on *HealthMindr*, a smartphone application targeting MSM that assesses PrEP eligibility monthly and allows users to order HIV tests, STI tests, and safe sex products. *HealthMindr* is currently under evaluation in a clinical trial.^21^ Based on estimates from the study protocol, we assumed that app users indicated for PrEP experienced a 4-fold higher probability of initiating PrEP while using the app. Given low engagement generally observed with STI mobile phone applications,^22^ we assumed that 20–25% of MSM were using the app at any given time for 3 months.

The adherence intervention was based on *Life-Steps for PrEP*, an in-clinic program that involves six counseling sessions over a 3-month period to discuss and address barriers to PrEP adherence.^23^ Based on pilot trial results, we modeled a 40% increase in the proportion of PrEP users on the intervention who were highly adherent, from 60% in the status quo to 87.1% for intervention participants. Based on HIV clinic capacity in Atlanta, we assumed that 30% of MSM initiating PrEP would engage in this intervention if offered.

The persistence intervention was based on *ePrEP* and *PrEP@Home* (both currently in trials), which provide a smartphone app, telemedicine visits with PrEP providers, and at-home HIV/STI screening to minimize clinic visits.^24,25^ Efficacy estimates from the *PrEP@Home* study protocol assume a 15.8% absolute increase in the proportion of PrEP users retained on PrEP after 1 year, which translated to a median PrEP duration of 347 days for intervention participants. Given the high acceptability of home-based PrEP care among MSM,^26^ we assumed that 40% of MSM starting PrEP would participate in this mode of PrEP delivery if offered.

### Intervention Strategies & Model Outcomes

A strategy consisted of one or more intervention options that decision makers could consider when choosing interventions to implement. We assessed a total of eight strategies: no continuum interventions (status quo), each intervention implemented alone, all combinations of two interventions, and a strategy that combines all three interventions.

Outcomes included infections averted, quality-adjusted life-years (QALYs), and direct health care costs. Individual QALYs varied by age, HIV disease stage, viral load, and treatment status. Individual costs varied by age, HIV status, PrEP use, and continuum intervention status (**Table 1**). Intervention costs were based on estimates of personnel time, overhead, direct medical, and resource/technology costs (Appendix, Section 11.6). All costs were reported in 2020 US dollars and were inflated using the consumer price index for medical care. As recommended by the Second Panel on Cost Effectiveness in Health and Medicine, costs and QALYs were discounted 3% annually.^6^ Broader societal impacts not captured explicitly in the model were summarized using an Impact Inventory (Appendix, Section 11.8).^6^

**Table 1.**
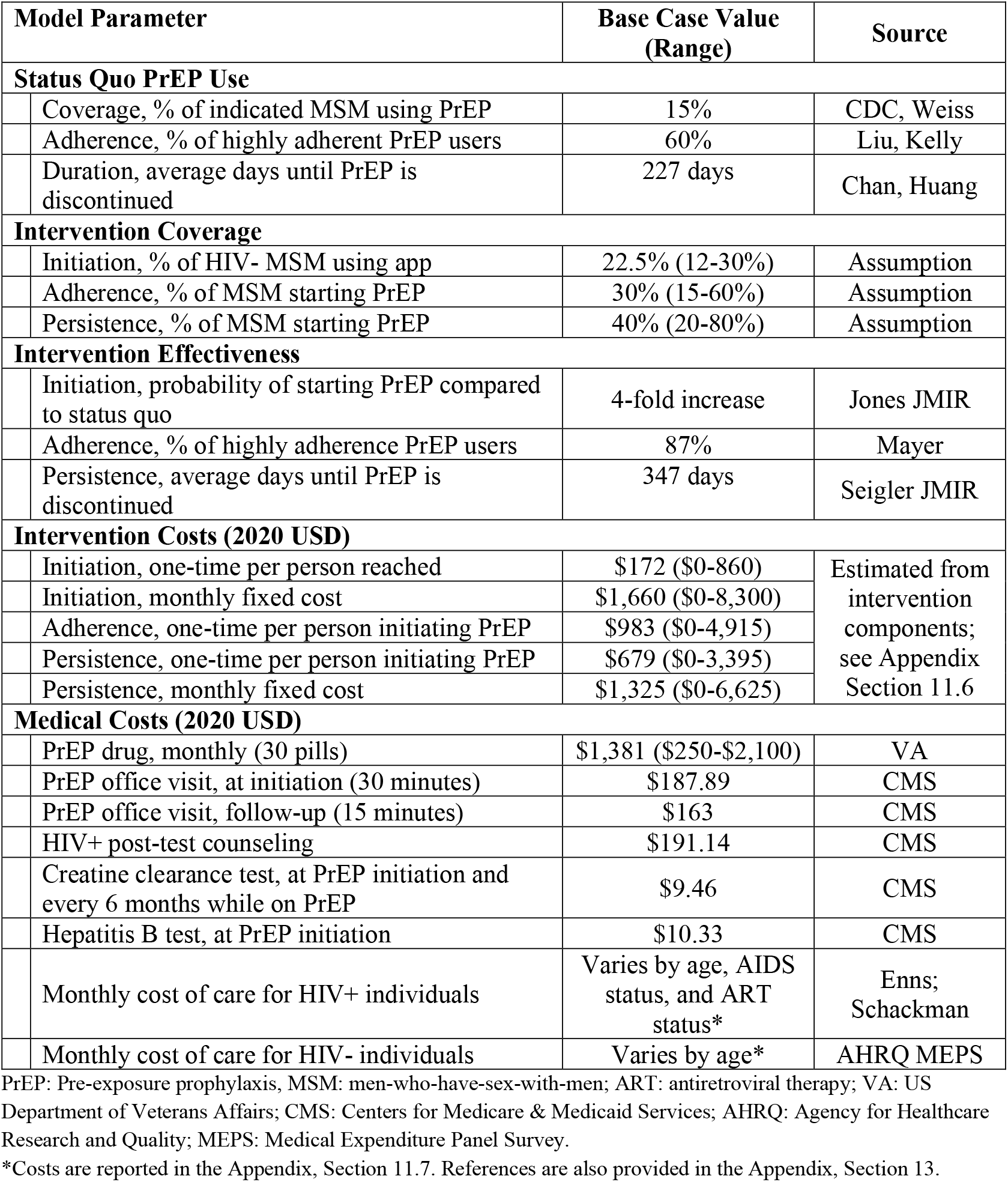
Model Input Values.

To estimate outcomes for each strategy, we simulated the population with relevant interventions implemented for the first 10 years of the time horizon (intervention period). After this point, the interventions were discontinued, with PrEP initiation, adherence, and persistence eventually returning to status quo levels. The simulation continued for an additional 10 years to capture residual downstream transmission benefits of the interventions. We then estimated the remaining lifetime costs and QALYs for individuals alive at the end of this total 20-year time horizon.

### Cost-Effectiveness Analysis

Lifetime costs and QALYs of the eight strategies were compared in a cost-effectiveness analysis where strategies were compared incrementally following best-practice guidelines.^6^ Strategies that cost more but were less effective (dominated) were eliminated as they would not be considered rational decisions. We then calculated incremental cost-effectiveness ratios (ICERs) by dividing incremental costs by the incremental benefits of moving from one strategy to the next best strategy. Strategies that gained benefits but less efficiently than a combination of two alternative strategies (weakly dominated) were also removed. A cost-effectiveness threshold of $100,000/QALY gained was used as a metric for cost-effectiveness when comparing non-dominated strategies. Uncertainty in ICERs was estimated from the 95% quantiles of bootstrapped distributions sampled from 10,000 simulation runs (Appendix, Section 12).

### Sensitivity Analyses

We varied PrEP costs, intervention costs, intervention coverage levels, and analysis perspective. The cost-effectiveness of PrEP medication influences the extent to which increasing person-time on PrEP is cost-effective. We therefore varied monthly PrEP costs between a highly subsidized cost ($250) up to the full cost of branded PrEP ($2,100), with generics falling somewhere in between ($800–$1,400). We varied intervention costs between 0– 5 times base case costs due to uncertainty in these parameters and to reflect potential differences in cost by geographic location. We also modeled low and high coverage scenarios to reflect different intervention scale-up levels. Lastly, we conducted the analysis from a payer perspective, where only intervention costs are included, as this would be of interest to potential funders.

## RESULTS

Compared to status quo PrEP use, strategies averted 44–973 (0.2–4.2%) new HIV infections over 20 years and gained between 141–7,383 (0.0045%–0.24%) discounted QALYs over a lifetime horizon in a population of 103,000 MSM (**Table 2**). The adherence intervention provided the smallest gains in QALYs and infections averted, while strategies that included the initiation intervention (I, IA, IP and IAP) achieved the largest QALYs gained and most infections averted. The highest PrEP coverage attained was 22.5% (compared to 15.1% in the status quo model), which occurred when initiation and persistence were used in combination; as expected, the adherence intervention alone had no impact on PrEP coverage.

**Table 2.** Clinical Outcomes, Costs, and Cost-Effectiveness for Men who Have Sex with Men (MSM) in Atlanta.

| Intervention | PrEP Coverage* (%) | Total Cost (2020 USD, billions) | PrEP Cost (% of Total) | Infections Averted (%) | Quality-Adjusted Life Years (QALYs) | Difference from Status Quo |  | Incremental Differences |  | ICER (Incremental cost/QALY gained) | ICER 95% Bootstrap CI (Incremental cost/QALY gained) |
| --- | --- | --- | --- | --- | --- | --- | --- | --- | --- | --- | --- |
|  |  |  |  |  |  | Cost (millions) | QALYs | Cost (millions) | QALYs |  |  |
| Status Quo | 15.1 | 45.74 | 5.31 | -- | 3,125,390 | -- | -- | -- | -- | -- | -- |
| Adherence (A) | 15.1 | 45.75 | 5.31 | 0.19 | 3,125,531 | 8.8 | 141 | 8.77 | 141 | 62,096 | (5,549, dominated)** |
| Initiation & Adherence (IA) | 20.0 | 46.27 | 6.17 | 3.14 | 3,131,575 | 534.2 | 6,185 | 525.4 | 6,044 | 86,927 | (82,824 , 91,391) |
| Combination of All (IAP) | 22.5 | 46.45 | 6.59 | 4.16 | 3,132,773 | 710.1 | 7,383 | 175.9 | 1,198 | 146,846 | (118,878 , 193,628) |
| Persistence (P) | 17.2 | 45.89 | 5.66 | 0.72 | 3,126,234 | 154.4 | 844 | -- | -- | ED | -- |
| Adherence & Persistence (AP) | 17.2 | 45.9 | 5.66 | 0.98 | 3,126,714 | 160.1 | 1,324 | -- | -- | ED | -- |
| Initiation (I) | 20.0 | 46.26 | 6.17 | 2.85 | 3,131,154 | 520.4 | 5,764 | -- | -- | ED | -- |
| Initiation & Persistence (IP) | 22.5 | 46.44 | 6.59 | 3.86 | 3,132,243 | 689.9 | 6,854 | -- | -- | ED | -- |

\* PrEP coverage is the percent of individuals indicated for PrEP who are using PrEP. The estimated population size of men-who-have-sex-with-men (MSM) in the Atlanta metropolitan area is 103,000 individuals.
\*\* Because the mean difference in QALYs between strategies “---” and “-A-“ was small in relation to the magnitude of stochastic variability in model outcomes, the bootstrapped confidence interval for this ICER was unreliable. For more detail on the uncertainty in this ICER refer to the Appendix Section 12.
USD: United States dollars; ICER: incremental cost-effectiveness ratio; ED: extended dominance, meaning the linear combination of two other strategies is more efficient than the labeled strategy; D: strong dominance, meaning the strategy produces fewer QALYs at a higher cost than another strategy.

There appeared to be minor synergistic impacts for combined interventions. For example, initiation combined with persistence resulted in slightly higher PrEP coverage than the sum of the coverage increases under each intervention implemented individually. Compared to the status quo, all strategies decreased healthcare spending, but these savings were largely outpaced by increased spending on PrEP medication, HIV screening, and interventions (**Figure 1**). For strategies involving initiation and persistence interventions (i.e., I, P, IP, and IAP), PrEP medication accounted for the largest increase in costs, which was 3–6 times greater than intervention costs.

**Figure 1.**
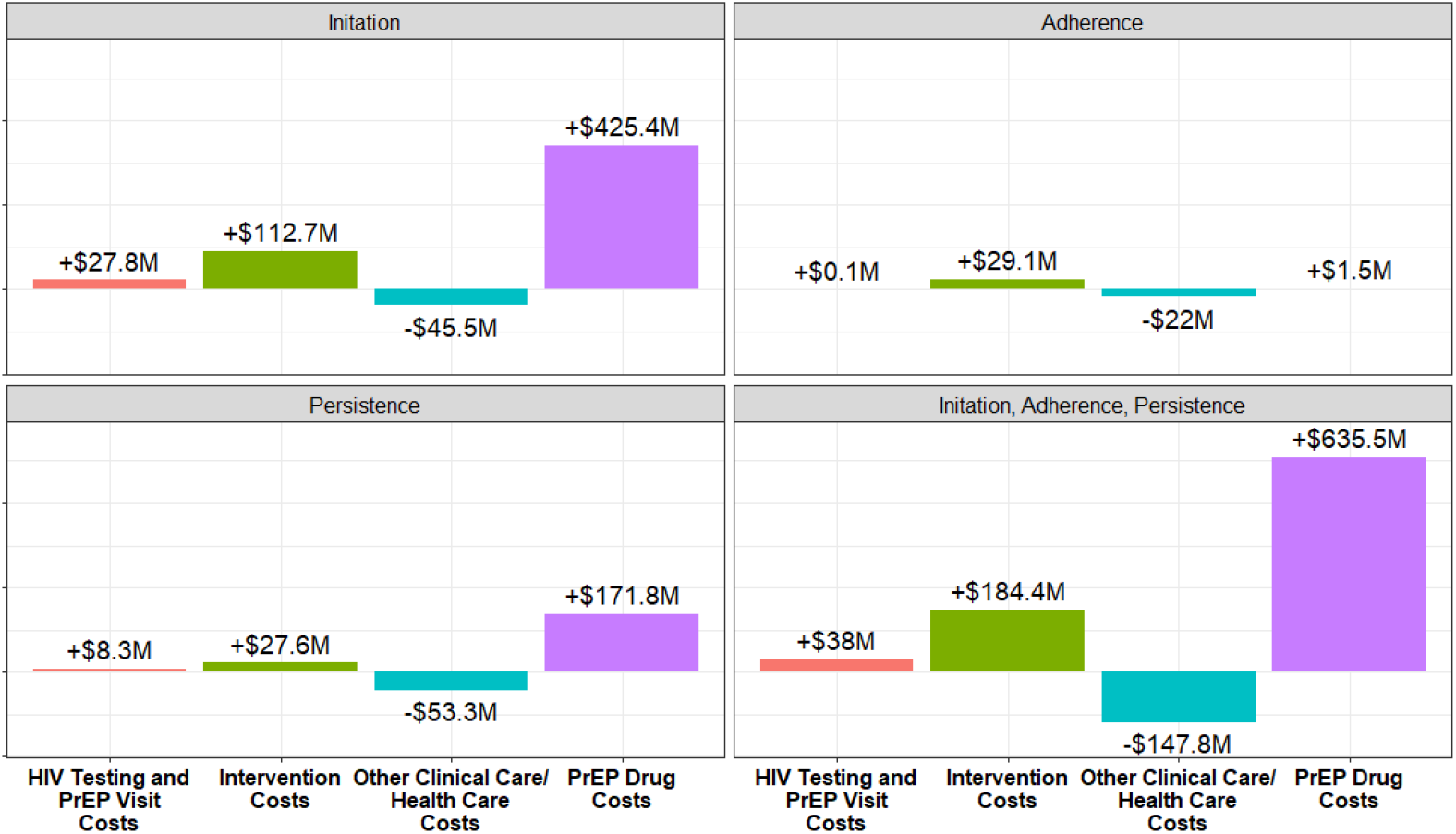
Cost Difference Relative to Status Quo. Higher spending on PrEP, HIV testing, and interventions results in lower downstream health care costs due to HIV infections averted (negative blue bars). **a)** When initiation is implemented alone, expenditure on the intervention is far outweighed by the increase in PrEP costs. **b)** When persistence is implemented, intervention costs ($2,760,000) are offset by savings in healthcare costs (-$5,800,000), but there are still substantial increases in PrEP costs. **c)** The adherence intervention results in inconsequential differences in HIV testing, PrEP visits, and PrEP drug costs compared to the status quo. Intervention spending is nearly offset by savings in healthcare costs. **d)** When all three interventions are combined, the largest savings in healthcare costs are achieved, but these savings are only 25% of the increase in spending on PrEP.

In comparing the costs and health benefits of the eight strategies, only three strategies were non-dominated: the adherence intervention alone, adherence and initiation combined, and the triple combination strategy of adherence, initiation, and persistence interventions. Together, these three strategies are considered efficient on the cost-effectiveness plane (**Figure 2**). The adherence intervention alone was the least costly but also least beneficial strategy and had an estimated ICER of $62,096 (95% CI: $5,549–Dominated) per QALY gained. Because the benefits conferred by the adherence intervention were so small, they were sometimes overwhelmed by random variation due to the stochastic nature of the model, leading to significant variation in the ICER that was not well summarized with a confidence interval (Appendix, Section 12). Combining the adherence and initiation interventions was the next least costly strategy on the efficient frontier, with an ICER of $86,927 (95% CI: $82,824–$91,391) per QALY gained relative to the adherence intervention alone. The most effective, but also most expensive strategy, was the combination of all three interventions, which had an ICER of $146,846 (95% CI: $118,878–$193,628) per QALY gained.

**Figure 2.**
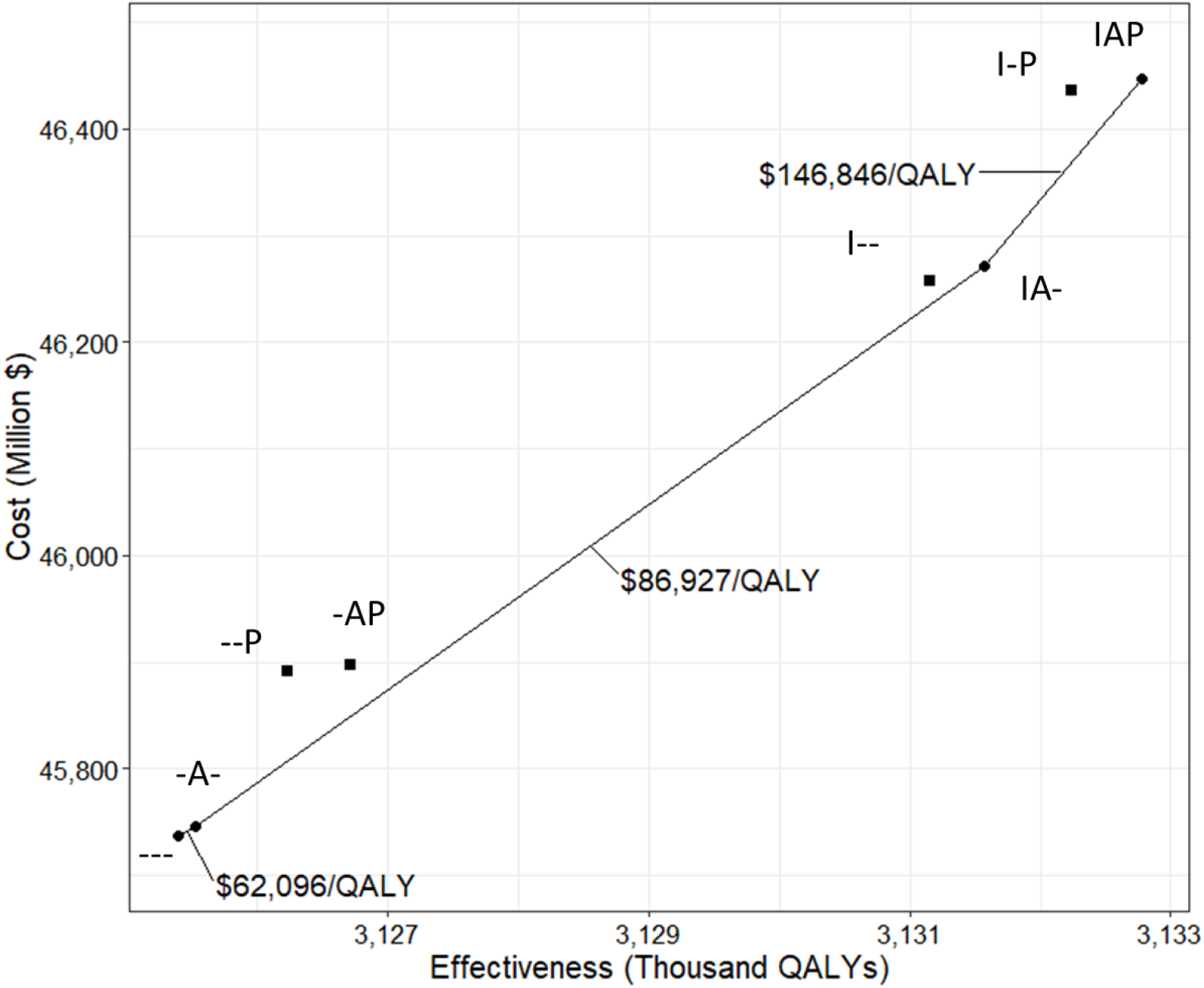
Cost-effectiveness plane. Each of the eight strategies are plotted according to their effectiveness (QALYs) and cost. Non-dominated strategies form the efficient frontier (line) and are the subset of strategies that would be considered by a rational decision maker. The cluster of strategies in the top right include initiation, while strategies clustered in the bottom left don’t include initiation. I = initiation, P = persistence, A = adherence.

At a willingness-to-pay threshold of $100,000/QALY gained, a strategy combining adherence and initiation interventions would be optimal; if a higher threshold of $150,000/QALY gained were used, the triple combination strategy would be preferred. All interventions were close to the efficient frontier so changes in base case assumptions may impact cost-effectiveness results. However, a consistent finding is that initiation results in the greatest QALY gains at a moderate increase in costs, while the addition of persistence results in a moderate increase in QALYs but a significantly higher cost per QALY gained.

At a willingness-to-pay threshold of $100,000/QALY gained, the triple combination strategy would be the optimal strategy if the monthly cost of PrEP was below $1,000 (**Figure 3a**). Sensitivity analyses also showed that cost-effectiveness was relatively dependent on intervention costs: if intervention costs were 1.6 times higher (60% increase) than their base case values, none of the strategies were cost-effective, and the status quo was preferred (**Figure 3b**). The adherence intervention was no longer cost-effective if its intervention costs were 1.8 times higher. Even if the persistence intervention costs were reduced to $0, the triple combination strategy still had an ICER >$100,000/QALY gained, and the combination of adherence and initiation interventions would be preferred.

**Figure 3.**
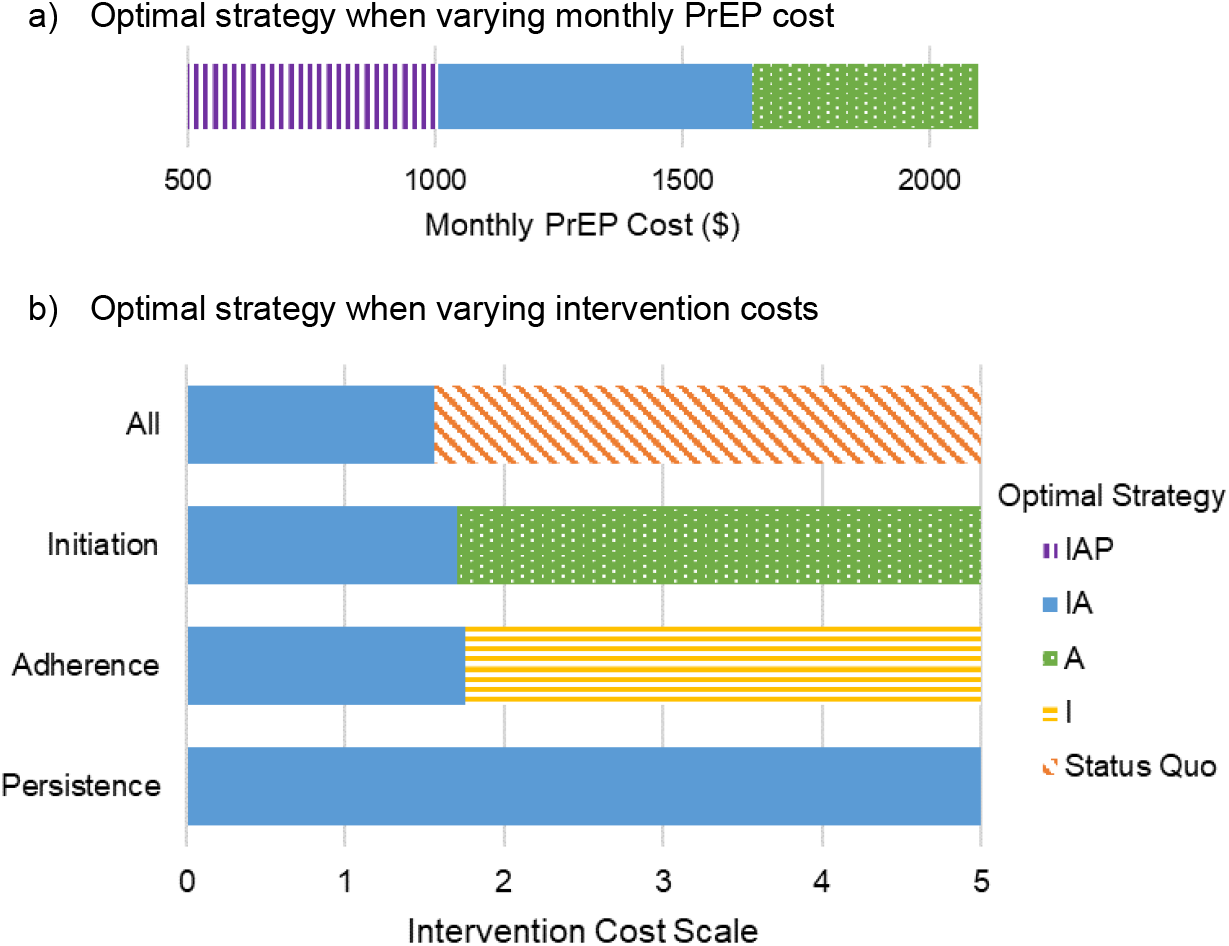
Sensitivity analyses: Optimal strategy using $100,000/QALY gained willingness-to-pay threshold. **a)** The cost of PrEP is varied from $250-$2,100/month on the x-axis and the optimal strategy is indicated by color and pattern. When PrEP costs less than $1,003, the triple combination strategy (initiation, adherence, and persistence) is the optimal cost-effective strategy. When PrEP costs increase above $1,644/month, adherence alone becomes the optimal strategy. **b)** Intervention costs are multiplied on a scale from 0 ($0 intervention costs) to 5 times base case costs. In the first row, all interventions are scaled simultaneously. When costs increase by 1.6 times (60%), no interventions are considered cost-effective and the status quo is the preferred strategy. In the second row, initiation costs are scaled while other intervention costs remain at base case levels. When initiation costs increase by 1.7 times (70%), adherence alone becomes the optimal strategy. In the third and fourth rows, adherence and persistence costs are scaled, respectively, while other intervention costs remain at base case levels. When adherence intervention costs increase by 1.8 times (80%), initiation alone becomes the optimal strategy. Even when the persistence intervention cost $0, the initiation & adherence combination strategy remains optimal. I = initiation, P = persistence, A = adherence.

Under low intervention coverage scenarios, strategies averted 0.1–1.7% infections and achieved a maximum of 18% PrEP coverage. Under high intervention coverage scenarios, strategies averted 0.4–7.2% infections and achieved a maximum of 28% PrEP coverage. The initiation and adherence combination strategy remained the optimal cost-effective option in both scenarios.

Using a payer perspective where only intervention costs were included, initiation alone, initiation combined with persistence, and the triple combination were the only non-dominated strategies. This differs from the healthcare perspective, where adherence interventions were typically non-dominated because they did not incur additional PrEP costs; when PrEP medication costs are not considered, initiation then persistence interventions achieve the same high QALY gains but with much lower costs, increasing their cost-effectiveness and dominating the less effective adherence intervention.

## DISCUSSION

In this study, we used cost-effectiveness analysis paired with a stochastic network-based HIV transmission model to estimate optimal strategies for investments in behavioral and clinical interventions that address gaps in the PrEP care continuum among MSM. We explicitly modeled real-world interventions that increase PrEP use and engagement. PrEP initiation was the primary intervention mechanism that averted infections, gained QALYs, and incurred additional costs. The adherence intervention did not achieve significant clinical benefit but was considered efficient because, under our assumptions, it did not increase PrEP costs (i.e., we assumed individuals incurred the full monthly cost of PrEP even if they had low adherence). Therefore, the small gains in QALYs that the adherence intervention achieved came at low cost compared to the persistence and initiation interventions, which increased PrEP costs by lengthening duration of PrEP use and getting more people to use PrEP, respectively. Real-world interventions to improve PrEP use can be cost-effective, but may achieve only limited gains in infections averted and overall PrEP coverage.

While cost-effective, interventions were also expensive. The initiation and adherence intervention strategy, with an ICER of $86,927/QALY gained, would be considered optimal using a willingness-to-pay threshold of $100,000/QALY gained, but would cost an estimated $151.2 million in intervention costs alone for our target population over the 10-year period (∼$15 million/year). The strategy combining initiation, adherence, and persistence was the most effective, yet also most expensive, costing an estimated $184.4 million in intervention costs with an ICER of $146,846/QALY gained. This combination strategy would be considered cost-effective using a $150,000/QALY gained threshold, which some suggest may be more appropriate for the US context.^27^

What strategy is considered optimal will also depend on the decision maker’s perspective. A related study performed a budget optimization analysis of strategies to minimize HIV incidence from a payer perspective, using similar PrEP interventions as this cost-effectiveness analysis.^15^ It found that investment in persistence interventions was optimal with lower budgets and investment in a combination of persistence and initiation, but *not* adherence, was optimal with higher budgets. Budget optimization analyses can inform a specific payer (e.g., CDC) how to best invest in interventions, yet they do not account for broader health system costs, impacts on quality of life, or overall efficiency of such investments relative to other health care spending. Results from our cost-effectiveness analysis differ from the budget optimization largely because cost-effectiveness accounts for increased spending on PrEP medications. For strategies that expanded PrEP coverage, spending on PrEP was 3–6 times more than spending on interventions. However, results from our sensitivity analysis using a payer perspective support the budget optimization finding that initiation and persistence are more efficient investments than adherence when PrEP and other healthcare costs are not considered.

Few studies have assessed the costs and projected impact of improving PrEP engagement in the US. One study by Chan *et al*. found that improving PrEP persistence had a moderate impact on preventing infections while improving adherence had no impact on new infections in Rhode Island.^11^ Our analysis supports this finding, with the additional key result that PrEP initiation had a much larger impact on preventing new infections than persistence. Our analysis also explicitly modeled the mechanisms by which improvements to persistence and adherence could be made, rather than assuming hypothetical increases in adherence and persistence.

Different assumptions about costs and intervention scale-up can yield very different cost-effectiveness estimates, even within the same geographic context. Krebs *et al*. found that expanded access to PrEP targeted to high-risk MSM in Atlanta averted 6% of new infections with an ICER of $6,123/QALY gained (2018 USD) compared to the status quo.^28^ In contrast, our initiation alone strategy averted 2.9% infections at $90,300/QALY gained when compared to the status quo (instead of incrementally with the other strategies). Krebs assumed lower costs for PrEP ($883/month vs. $1,381/month) and lower one-time intervention costs ($18/person vs. $172/person). Krebs also assumed a higher PrEP expansion rate based on historic increases in average PrEP use between 2015-2017. After 18 months of intervention implementation, it appears the resulting PrEP coverage remained constant without interventions or additional costs. In contrast, our analysis assumed intervention effectiveness based on clinical trial estimates and intervention implementation for 10 years with PrEP coverage eventually returning to status quo levels when interventions ended. Low intervention costs and effectiveness estimates based on historic rates may not be realistic moving forward as we work towards engaging harder-to-reach populations in PrEP.

Results of our study found interventions could be cost-effective, but also that interventions were costly and did not come close to achieving EHE targets given reasonable assumptions regarding intervention participation. Even when all three interventions were used in combination, the maximum PrEP coverage achieved was only 23% in the base case and 28% in the high intervention coverage scenario. While these are substantial increases from baseline levels of 15%, they are still well-below national targets to achieve 50% of people with indications using PrEP by 2025.^29^ This highlights how much time, effort, and resources may be needed to engage people who face significant barriers to PrEP use. Continued expansion of PrEP coverage towards increasingly “hard-to-reach” populations will likely involve multiple interventions that may require increased investment and intensity.^4,30^ Current accepted thresholds of cost-effectiveness ($100,000-$150,000/QALY gained) in the US may not be realistic if we are truly aiming to end HIV. The recent availability of generic PrEP may help make investment in PrEP interventions more cost-effective, particularly if the price of generics decreases as more competitors enter the market. However, more systemic changes to improve healthcare access and to lower costs of care for vulnerable populations are likely still needed.

## Limitations

First, two interventions we selected are currently in clinical trials, so our effectiveness estimates were based on assumed effect sizes within protocol power calculations. While power calculations are typically conservative, trial outcomes will nonetheless provide better informed effectiveness estimates, with the caveat that trial effectiveness can still be different from effectiveness in practice. Second, we used Atlanta-specific demographic, epidemic, and cost data whenever possible. Results may be generalizable to other metropolitan areas with similar characteristics (such as similar HIV incidence rates and PrEP coverage levels, but generalizability may be limited for other contexts. Third, due to computational burden we did not conduct more extensive sensitivity analyses. While we estimated 95% confidence intervals to reflect model uncertainty given our assumptions, further assessments such as varying intervention effectiveness and conducting probabilistic analyses would be beneficial. Future research will seek to address these limitations. Fourth, we did not directly analyze racial, ethnic, or socioeconomic disparities in healthcare access and HIV risk, despite their well-documented existence and importance in PrEP access and use.^3,17,29^ Sexual network and epidemic parameters were stratified by race/ethnicity (Black, Hispanic, white) to capture Atlanta-specific characteristics, but QALYs and interventions were applied equally across racial and ethnic groups. We did not investigate options for prioritizing interventions to specific high-risk or disproportionately affected groups, such as young Black or Hispanic MSM. Future work could investigate these strategies since both EHE and the US National Strategic Plan include goals to address disparities;^1,29^ however, particular attention would need to be paid to ethical concerns when using cost-effectiveness to inform resource prioritization between population subgroups.^6^

## Conclusions

Our analysis highlights the costs and benefits of three novel interventions targeting the PrEP continuum of care. Strategies that included the initiation intervention achieved higher PrEP coverage, higher infections averted, and more QALYs gained than strategies that included only adherence and/or persistence. However, our analysis also showed that even with substantial investment in PrEP uptake and engagement interventions, only limited gains were made in expanding PrEP coverage and reducing HIV incidence. Additional interventions and investments would be needed to reach *Ending the HIV Epidemic* goals.

## Data Availability

Model code used in the analysis is publicly available on Github.

https://github.com/EpiModel/PrEP-CEA

https://github.com/statnet/EpiModelHIV

## ACKNOWLEDGEMENTS

We’d like to thank Aaron Siegler, Jeb Jones, and Patrick Sullivan for providing cost estimates for the *HealthMindr, ePrEP*, and *PrEP@Home* interventions. This work was supported by US National Institutes of Health grant R01 AI138783.

