## Supplemental Appendix for "Cost-Effectiveness of Interventions to Improve HIV Pre-Exposure Prophylaxis Initiation, Adherence, and Persistence among Men Who Have Sex with Men"

#### Supplementary Appendix

Model details are in Sections 1-10 and analysis methods with additional cost-effectiveness results are in Sections 11-13.

### 1 MODEL INTRODUCTION

The first ten sections of this supplementary technical appendix describe the mathematical model structure, parameterization, and statistical analysis of the accompanying paper in further detail.

#### 1.1 Model Framework

The mathematical models for HIV transmission dynamics presented in this study are network-based transmission models in which uniquely identifiable sexual partnership dyads were simulated and tracked over time. This partnership structure is represented through the use of temporal exponential-family random graph models (TERGMs), described in Section 3. On top of this dynamic network simulation, the epidemic model represents demography (entries, exits, and aging), interhost epidemiology (disease transmission), intrahost epidemiology (disease progression), and clinical epidemiology (disease diagnosis and treatment and prevention interventions). Individual attributes related to these processes are stored and updated in discrete time over the course of each epidemic simulation.

The modeling methods presented here utilize and extend the *EpiModel* software platform to incorporate HIV-specific epidemiology and transmission dynamics. The HIV extensions for men who have sex with men (MSM) were originally developed by Goodreau et al. for use in prior modeling studies of MSM in the United States and South America,[1–3] and subsequently used to model HIV preexposure prophylaxis (PrEP) among US MSM.[4–7] The most recent innovation in our modeling platform has been to incorporate primary data from the ARTnet study of MSM in the United States directly into the workflow for parameterizing the network and behavioral components.[8]

#### 1.2 Model Software

The models in this study were programmed in the R and C++ software languages using the *EpiModel* [http://epimodel.org/] software platform for epidemic modeling. *EpiModel* was developed by the authors for simulating complex network-based mathematical models of infectious diseases, with a primary focus on HIV and sexually transmitted infections (STIs).[9] *EpiModel* depends on *Statnet* [http://statnet.org/], a suite of software in R for the representation, visualization, and statistical analysis of complex network data.[10]

*EpiModel* allows for a modular expansion of its built-in modeling tools to address novel research questions. We have developed a set of extension modules into a software package called *EpiModelHIV*. This software is available for download, along with the scripts used in the execution of these models. The tools and scripts to run these models are contained in two GitHub repositories:

- [http://github.com/statnet/EpiModelHIV] contains the general extension software package. Installing this using the instructions listed at the repository homepage will also load in *EpiModel* and the other dependencies. We use a branching repository architecture on Github; the branch of the repository associated with this research project is *PrEP-CEA*.
- [http://github.com/EpiModel/PrEP-CEA] contains the scripts to execute the models and to run the statistical analyses provided in the manuscript.

##### 1.3 Core Model Specifications

We started with a network size of 10,000 MSM aged 15 to 65 to represent the larger population of sexually active MSM in the Atlanta metropolitan area. The population size was allowed to increase and decrease over time with arrivals into the sexually active population at age 15 and departures related to mortality or aging out of the sexually active population at age 65. MSM were stratified by black, Hispanic, and white/other (hereafter in the text, called white) race/ethnicity in proportions equivalent to Census-derived proportions. Further details on the demography (race and age) are provided in Section 5. We used a three-stage simulation framework, first calibrating the model to diagnosed HIV prevalence and HIV care continuum parameters for 60 years of burn-in time (Stage 1), then calibrating the model to current estimated levels of PrEP coverage for 5 years of burn-in time (Stage 2), and then simulating the reference and counterfactual intervention scenarios for 10 years in intervention scenarios (Stage 3). The time unit used throughout the simulations was one week. Unless otherwise noted, all rate-based parameters listed below are to be interpreted as the rate per week and all duration-based estimates are to be interpreted as the duration in weeks.

#### 2 THE ARTnet STUDY

This model featured an innovative parameterization design in which primary individual-level and partnership-level data were used to fit statistical models for summary statistics that were then entered into the epidemic model. The primary data source for network structure and behavioral data was the ARTnet study, described below. Wherever possible, we used primary data from this study for model parameterization, and only relied on the secondary published literature for model parameters that could be generalized across target populations (e.g., HIV natural history or clinical response parameters).

##### 2.1 Study Design

This analysis used data collected in the ARTnet study of MSM in the United States in 2017–2019.[8] MSM were recruited directly after participating in the American Men's Internet Study (AMIS),[11] a parent web-based study about MSM sexual health that recruited through banner ads placed on websites or social network applications. At the completion of AMIS, MSM were asked to participate in ARTnet, which focused on sexual network features. ARTnet data collection occurred in two waves (following AMIS): July 2017 to February 2018 and September 2018 to January 2019.

Eligibility criteria for ARTnet were male sex at birth, current male cisgender identity, lifetime history of sexual activity with another man, and age between 15 and 65. Respondents were deduplicated within and across survey waves (based on IP and email addresses), resulting in a final sample of 4904 participants who reported on 16198 sexual partnerships. The Emory University Institutional Review Board approved the study.

##### 2.2 Primary Measures

ARTnet participants were first asked about demographic and health-related information. Covariates used in this analysis included race, age, ZIP Code of residence, and current HIV status. ZIP Codes were transformed into Census regions/divisions and urbanicity levels by matching against county databases (using standardized methods for selecting county in the small number of cases when ZIP Codes crossed county lines). Participants reporting as never testing for HIV, having indeterminate test results, or never receiving test results were classified as having an unknown HIV status.

Participants were then asked detailed partner-specific questions for up to their most recent 5 partners. The detailed partner-specific questions included attributes of the partner and details about the partnership itself. Partner attributes considered here included age, race/ethnicity, and HIV status. Participants were allowed to report any partner attribute as unknown. When partner age was unknown, age was imputed based on a response to a categorical question (e.g., 5–10 years younger/older, 2–5 years younger/older). Partnerships were classified into three types: main (respondent reported they considered this partner a "boyfriend, significant other, or life partner") casual (someone they have had sex with more than once, but not a main partner), and one-time.[12] For one-time partners, we asked for the date that sexual activity occurred. For persistent (main and casual) partnerships, we asked for the date of most recent sex, the date of first sex (which could have been prior to the past year), and whether the partnership was ongoing (if the participant expected sexual activity would occur in the future). For each partnership, we asked whether (for one-time) or how frequently (for persistent) anal sex occurred.

Outcome measures include descriptive statistics for characteristics of participants and their reported partnerships, and the aggregate network statistics used to fit the TERGMs underlying epidemic simulations on dynamic networks. The network statistics include ego degree, attribute mixing in partnerships, and the current length of ongoing partnerships, stratified by the attributes of persons and partnerships. Degree is a property of individuals, whereas mixing and length are properties of partnerships. Degree was defined as the ongoing number of persistent partners measured on the day of the survey (includes main and casual partnerships). Degree is not defined for one-time partnerships, so for these we instead calculated a weekly rate of new contacts by subtracting the total main and casual partners from the total past-year partners and dividing by 52. Partnership length for ongoing main and casual partnerships was calculated by taking the difference between the survey date and the partnership start date. The mean length of ongoing partnerships is the network statistic needed for TERGM estimation; the logic and derivation are explained here.[9] Mixing was measured by the relative frequency of partnerships that occurred within and between groups defined by race/ethnicity, and age.

##### 2.3 Statistical Analysis

We fit a series of generalized linear models (GLMs) to estimate summary statistics for features of the sexual network structure and the behavior within partnerships. Specific GLM parameterizations are detailed below in the discussion of each set of model parameters. Common across all models was the general approach of including geography of residence as a main effect with two levels (Atlanta versus all other areas). This allowed for the model coefficients and predicted summary statistics to vary by geography while ensuring stability of outcomes under the assumption of conditional exchangeability.

#### 3 NETWORKS OF SEXUAL PARTNERSHIPS

We modeled networks of three interacting types of sexual relations: main partnerships, casual (but persistent) partnerships, and one-time anal intercourse contacts. We first describe the methods conceptually, including the parameters used to guide the model and their derivation, and then present the formal statistical modeling methods. Consistent with our parameter derivations, all relationships are defined as those in which anal intercourse is expected to occur at least once.

##### 3.1 Conceptual Representation of Sexual Networks

Our modeling methods aim to preserve certain features of the cross-sectional and dynamic network structure as observed in our primary data, while also allowing for mean relational durations to be targeted to those reported for different groups and relational types. Our methods do so within the context of changing population size (due to births, deaths, arrivals and departures from the population) and changing composition by attributes such as age. The broader motivation, methodological details, and link between models and primary data are described here.[9]

The network features that we aim to preserve are as follows:

- Persistent (Main and Casual) Partnerships
    - The mean degree (number of ongoing partners), stratified by main and casual partnership types, and the proportion of men with concurrency (2 or more ongoing partners) for each partnership type, at any time point.
    - Variations in the mean degree specific to each persistent partnership type by:
        - Race/ethnicity group (3 categories for black, Hispanic, and white MSM).

        - Age group (5 categories for 15–24, 25–34, 35–44, 45–54, and 55–64).
        - Cross-type degree: Degree in the other persistent partnership type (e.g., mean degree of MSM for main partnerships given current casual degree of 0, 1, 2, 3).
    - Selection of partners within the same race/ethnicity group (mixing by race/ethnicity).
    - Selection of partners within the same age group (mixing by age).
    - Mean partnership durations, stratified by main and casual partnership types, and by mixing within age groups.
- One-Time Partnerships
    - The overall rate of having one-time anal intercourse partnerships per week.
    - Variations in this contact rate by:
        - Race/ethnicity group.
        - Age group.
        - Total persistent degree (sum of main and casual partnerships ongoing).
        - Risk level heterogeneity above and beyond the risk heterogeneity associated with demographics (mean partnership rates for five quintiles of MSM stratified by mean one-time rates).
    - Selection of partners within the same race/ethnicity group (mixing by race/ethnicity).
    - Selection of partners within the same age group (mixing by age).
- Common to Persistent and One-Time Partnership Types
    - Prohibitions against MSM with incompatible sexual positioning roles (e.g., no partnerships between exclusively receptive MSM).

##### *3.1.1 Overall Mean Degree for Persistent Partnerships*

Ongoing persistent partnerships (whether main or casual) were defined from the partnership-level ARTnet dataset as those in which sex had already occurred more than once, and in which the respondent anticipated having sex again. The momentary main or casual mean degree is then defined as the mean of the degree of all MSM for main or casual partnerships on the day of study. We estimated this with a Poisson model with main or casual degree as the outcome and a dummy variable for Atlanta residence as the predictor and then exponentiating the coefficients, resulting in an estimated mean main degree of 0.396 and a mean casual degree of 0.541.

In addition, we modeled the proportion of MSM with concurrency (degree of 2 or more) by partnership type. This was estimated with logistic regression models for binary outcomes with a dummy variable for Atlanta residence as the predictor. Taking the inverse of the logit of the coefficient yielded the predicted probabilities of 0.9% for main concurrency and 14.5% for casual concurrency.

##### *3.1.2 Heterogeneity in Mean Degrees for Persistent Partnerships*

We estimated the heterogeneity in main and casual mean degree by fitting three Poisson regression models. For race/ethnicity, we estimated the mean degree for each group within the target population by including dummy variables for city and race/ethnicity. For age, we modeled the non-linear relationship between age and mean degrees by including city, age group, and square root of age group to allow for a non-linear relationship between age and the outcome. For cross type degree, we modeled the mean degree for main partnerships as a function of degree of casual partnerships, and vice versa, again with city also as a predictor. For each of the 6 models (2 partnership types times three predictors of interest), we fit the statistical models and then exponentiated the coefficients to obtain the rates for each stratum. Those are shown in Supplemental Table 1 below.

| **Supplemental Table 1.** Heterogeneity in Mean Main and Casual Degree by Race/Ethnicity, Age Group, and Cross Type Degree of Ego (Respondent) | | |
|---|---|---|
| **Predictor** | **Main Mean Degree** | **Casual Mean Degree** |
| **Race/Ethnicity** | | |
| Black | 0.566 | 0.605 |
| Hispanic | 0.470 | 0.513 |
| White | 0.823 | 0.534 |
| **Age Group** | | |
| 15–24 | 0.795 | 0.297 |
| 25–34 | 0.697 | 0.479 |
| 35–44 | 0.577 | 0.615 |
| 45–54 | 0.448 | 0.701 |
| 55–64 | 0.326 | 0.742 |
| **Cross Type Degree** | | |
| 0 | 0.440 | 0.614 |
| 1 | 0.352 | 0.377 |
| 2 | 0.282 | 0.009 |
| 3 | 0.225 | — |

###### *3.1.3 Mixing by Race/Ethnicity and Age for Persistent Partnerships*

Respondents reported on their perception of the race and ethnicity (Hispanic/non-Hispanic) for each partner. We categorized the respondents' and partners' races into three mutually exclusive groups: black, Hispanic, and white. Using logistic regression models, we estimated the proportion of partnerships that were between MSM of the same race (within-group mixing) by evaluating relationship between the respondent group and partner group as a binary outcome (using the geography of residence predictor as a main effect with two levels, Atlanta versus all other areas). The inverse logit of the coefficients is then interpreted as the predicted probability of a same-race/ethnicity partnership. The values were 76.5% for main partnerships and 63.3% for casual partnerships.

For mixing by age, we used a model parameterization for the 5-category age group that allowed for differences in the level of age mixing that could vary by age group (differential homophily). We fit a logistic regression model for partnerships, with being in a partnership of the same age group as the outcome and the age group of the respondent as the main predictor. With the inverse logit transformation, the probabilities of partnerships within the same age group, stratified by partnership type are shown in Supplemental Table 2 below.

| **Supplemental Table 2.** Proportion of Main and Casual Partnerships within the Same Age Group, by Age of Ego (Respondent) | | |
|---|---|---|
| **Age Group** | **Main Within Group** | **Casual Within Group** |
| 15–24 | 79.5% | 56.4% |
| 25–34 | 69.7% | 43.8% |
| 35–44 | 57.8% | 31.9% |

| | | |
|---|---|---|
| 45–54 | 44.8% | 22.1% |
| 55–64 | 32.6% | 14.6% |

##### *3.1.4 Duration of Persistent Partnerships*

We model partnership dissolution as a heterogenous, geometrically distributed process with unique parameters for each relational type. The geometric distribution for relational durations implies a “memoryless process,” which is a common assumption within ordinary differential equation modeling. Although this assumption implies that the rate of dissolution does not depend on the current age of the partnership, the overall exponential shape of the dissolution distribution matches reasonably well to empirical data on relational durations. The fit is improved considerably when the partnership types are stratified, as we do here, implying a mixture of geometric distributions. Once one-time contacts are removed, and longer-duration main partnerships are separated from shorter-term causal partnerships, the set of geometric distribution fits the empirical data on partnership durations well.

The fit is improved further by stratifying based on the interaction between partnership type and the age of both members within the dyad. For this analysis, we explored how relationship duration varied by multiple demographic characteristics, and unsurprisingly age was most strongly associated with duration. For this model parameterization, we specifically elected to estimate and input parameters based on matched age groups (that is, partnerships between two persons of the same age).

As detailed in previous work,[1,9] for memoryless processes, the expected age of an extant (ongoing) relationship at any moment in time is an unbiased estimator of the expected uncensored duration of relationships, given the balancing effects of right-censoring and length bias for this distribution. Raw relational ages were calculated as the difference between first sex date and the study date for each dyad the ego reported sex with more than once in the interval. To derive our estimator of relational age, we take the median of the observed distribution and then calculate the mean for the geometric distributions associated with that median. To account for estimation within the Atlanta target population, we weighted this estimator by the inverse of the relative differences in Atlanta partnerships to non-Atlanta partnerships.

The resulting expected relational ages are summarized in Supplemental Table 3 below.

**Supplemental Table 3.** Duration of Main and Casual Partnerships by Dyadic Age Group of Ego (Respondent) and Alter (Partner)

| **Dyadic Age Group** | **Main Relational Age (Weeks)** | **Casual Relational Age (Weeks)** |
|---|---|---|
| Both 15–24 | 71.2 | 50.5 |
| Both 25–34 | 253.5 | 72.5 |
| Both 35–44 | 523.3 | 112.1 |
| Both 45–54 | 637.1 | 161.3 |
| Both 55–64 | 903.1 | 147.4 |
| Different Groups | 217.9 | 106.4 |

##### *3.1.6 Overall Mean One-Time Contact Rate*

In addition to persistent main and casual partnerships, we modeled one-time sexual contacts involving anal intercourse based on ARTnet reports on the number and variation in these types of relations. As noted above, degree is not defined for one-time contacts, so for these we instead calculated a weekly rate of new contacts by subtracting the total main and casual partners from the total past-year partners. We estimated the weekly rate by fitting a Poisson regression model with the count of one-time contacts as a function of city, exponentiating the coefficient to get the predicted count, and dividing by 52 to get the week rate. The overall mean one-time contact rate was 0.076 anal intercourse contacts per week.

##### 3.1.7 *Heterogeneity in One-Time Contact Rates*

Heterogeneity in one-time contact rates was modeled with four Poisson regression models to estimate the rates as a function of race/ethnicity, age group, risk level strata, and total persistent (main plus casual) degree. Similar to the one-time rate, we fit these models with geography of residence as a main effect (which had two levels, Atlanta versus all other areas, with the former level used for predictions) and exponentiated the coefficients and then divided by 52 to get the group-specific rates. For age group, similar to the estimation of degree, we modeled this non-linearly by including age group and the square root of age group as the joint predictors (along with city). The results are shown in Supplemental Table 4 below.

**Supplemental Table 4.** Weekly One-Time Contact Rates by Race/Ethnicity, Age Group, Risk Level, and Total Persistent Degree of Ego (Respondent)

| Predictor | Weekly Contact Rate |
|---|---|
| **Race/Ethnicity** | |
| Black | 0.062 |
| Hispanic | 0.071 |
| White | 0.079 |
| **Age Group** | |
| 15–24 | 0.048 |
| 25–34 | 0.075 |
| 35–44 | 0.089 |
| 45–54 | 0.093 |
| 55–64 | 0.087 |
| **Risk Level Quintile** | |
| 1 | 0.000 |
| 2 | 0.000 |
| 3 | 0.012 |
| 4 | 0.043 |
| 5 | 0.326 |
| **Total Persistent Degree** | |
| 0 | 0.049 |
| 1 | 0.057 |
| 2 | 0.121 |
| 3+ | 0.284 |

##### 3.1.8 *Mixing by Race/Ethnicity and Age for One-Time Contacts*

We used a similar approach to within-group mixing by race/ethnicity and age group for one-time contacts to the one used for persistent contacts, with one difference that we did not model differential homophily by age group to improve model stability. Therefore, the overall proportion of one-time contacts that were within the same race/ethnic group was 67.6% and the proportion of one-time contacts that were within the same age group was 32.8%.

##### 3.1.9 *Mixing by Sexual Role Across All Partnership Types*

We assign men a fixed sexual role preference (exclusively insertive, exclusively receptive, versatile). The model then includes an absolute prohibition, such that two exclusively insertive men cannot partner, nor can two exclusively receptive men. We estimated the proportion of men that were in each category (insertive, receptive, and versatile) by analyzing whether men had only insertive anal intercourse, only receptive anal intercourse, or both insertive and receptive anal intercourse (respectively) in their past five anal partnerships over the past year. These proportions were stratified (restricted) by geography of residence to the city of Atlanta. The proportions were: 18.5% exclusively insertive, 27.1% exclusively receptive, and 54.4% versatile.

#### 3.2 Statistical Representation of Sexual Networks

Exponential-family random graph models (ERGMs) and their dynamic extension temporal ERGMs (TERGMs) provide a foundation for statistically principled simulation of local and global network structure given a set of target statistics from empirical data. Main and casual relationships were modeled using TERGMs,[13] since they persist for multiple time steps. One-time contacts, on the other hand, were modeled using cross-sectional ERGMs.[14] Formally, our statistical models for relational dynamics can be represented as five equations for the conditional log odds (logits) of relational formation and persistence at time $t$ (for main and casual relationships) or for relational existence at time $t$ (for one-time contacts):

$$logit\left(P\left(Y_{ij,t}=1 \middle| Y_{ij,t-1}=0, Y_{ij,t}^{C}\right)\right) = \theta_m^{+\prime}\partial\left(g_m^{+}(y)\right) \quad \text{Main partnership formation}$$

$$logit\left(P\left(Y_{ij,t}=1 \middle| Y_{ij,t-1}=0, Y_{ij,t}^{C}\right)\right) = \theta_c^{+\prime}\partial\left(g_c^{+}(y)\right) \quad \text{Casual partnership formation}$$

$$logit\left(P\left(Y_{ij,t}=1 \middle| Y_{ij,t-1}=1, Y_{ij,t}^{C}\right)\right) = \theta_m^{-\prime}\partial\left(g_m^{-}(y)\right) \quad \text{Main partnership persistence}$$

$$logit\left(P\left(Y_{ij,t}=1 \middle| Y_{ij,t-1}=1, Y_{ij,t}^{C}\right)\right) = \theta_c^{-\prime}\partial\left(g_c^{-}(y)\right) \quad \text{Casual partnership persistence}$$

$$logit\left(P\left(Y_{ij,t}=1 \middle| Y_{ij,t}^{C}\right)\right) = \theta_o{}'\partial\left(g_o(y)\right) \quad \text{One-time contact existence}$$

where:

- $Y_{ij,t}$ = the relational status of persons $i$ and $j$ at time $t$ (1 = in relationship/contact, 0 = not).
- $Y_{ij,t}^{C}$ = the network complement of $i,j$ at time $t$, i.e. all relations in the network other than $i,j$.
- $g(y)$ = vector of network statistics in each model (the empirical statistics defined in the tables above).
- $\partial\left(g(y)\right)$ = the change in $g(y)$ when $Y_{ij}$ is toggled from 0 to 1 (for formation models) or 1 to 0 (for persistence models).
- $\theta$ = vector of parameters in the model.

For $g(y)$ and $\theta$, the superscript distinguishes the formation model (+), persistence model (-) and existence models (neither). The subscript indicates the main (m), casual (c) and one-time (o) models.

The recursive dependence among the relationships renders the model impossible to evaluate using standard techniques; we use MCMC in order to obtain the maximum likelihood estimates for the $\boldsymbol{\theta}$ vectors given the $\boldsymbol{g}(\boldsymbol{y})$ vectors.

Our method of converting the statistics laid out in Section 3.1 into our fully specified network models consists of the following steps:

1. Construct a cross-sectional network of 10,000 men with no relationships.
2. Assign men demographics (race/ethnicity and age) based on Census data for Atlanta and assign men sexual roles based on frequencies listed above, as well as one-time risk quintiles (20% of the men in each race per quintile).
3. Calculate the target statistics (i.e., the expected count of each statistic at any given moment in time) associated with the terms in the formation model (for the main and casual partnerships) and in the existence model (for one-time contacts).
4. Assign each node a place-holder main and casual degree (number of ongoing partnerships) that is consistent with the estimated distributions and store these numbers as a nodal attribute. (Note: this does not actually require individuals to be paired up into the partnerships represented by those degrees).
5. For the main and casual networks, use the mean relational durations by age group combination to calculate the parameters of the persistence model, using closed-form solutions, given that the models are dyadic-independent (each relationship's persistence probability is independent of all others).
6. For the main and casual networks, estimate the coefficients for the formation model that represent the maximum likelihood estimates for the expected cross-sectional network structure.
7. For the one-time network, estimate the coefficients for the existence model that represent the maximum likelihood estimates for the expected cross-sectional network structure.

Steps 5–7 occur within the *EpiModel* software and use the ERGM and STERGM methods therein. They are completed efficiently by the use of an approximation in Step 6.[15] During the subsequent model simulation, we use the method of Krivitsky[16] to adjust the coefficient for the edges term in each model at each time step, in order to preserve the same expected mean degree (relationships per person) over time in the face of changing network size and nodal composition. At all stages of the project, simulated partnership networks were checked to ensure that they indeed retained the expected cross-sectional structure and relational durations throughout the simulations.

#### 4 BEHAVIOR WITHIN SEXUAL PARTNERSHIPS

In this study, we model three phenomena consecutively within relationships at each time step: the number of anal intercourse sex acts, condom use per sex act, and sexual role per sex act. We simulate these within all relationships regardless of HIV status (whether diagnosed or not).

##### 4.1 Anal Intercourse Acts Per Partnership

The rate of anal intercourse is applicable to persistent (main and casual) partnerships in which there are repeated anal intercourse acts between the start and end of the partnership. We use ARTnet data for the overall rate and predictors of variation in rates unique to each partnership type. For one-time contacts, we assumed that the number of anal intercourse exposures was one by definition, although there could have been multiple anal intercourse acts within an exposure due to role versatility (see Section 4.4). The modeling of act rates here is based on the expectation that changes in coital frequency depend on race/ethnicity, age, diagnosed HIV status, and partnership type.

###### *4.1.1 Measurement of Acts in ARTnet*

We measured the number of acts within each reported partnership within the ARTnet study by asking participants about the frequency of anal intercourse acts. Study participants could report on the average number of acts within the partnership over the past year by week, month, year, or total partnership duration. We then scaled this into a total weekly act rate. The final ARTnet partnership-level dataset on 16198 partnerships includes this weekly rate as the outcome and predictors at the individual and dyadic level that we used for statistical modeling as described below.

###### *4.1.2 Statistical Models of Act Rates*

With this partnership-level dataset, we then modeled the count of acts per year per partnership based on the Poisson regression formula:

$$Y_i \sim \beta_0 + \beta_1 X_1 + \beta_2 X_1^2 + \beta_3 X_2 + \beta_4 X_3 + \beta_5 X_1 X_3 + \beta_6 X_4 + \beta_7 X_4^2 + \beta_8 X_5 + \beta_9 X_6$$

where:

$Y_i$ = Log of the count of acts per year.

$X_1$ = Duration of partnership in weeks at the survey date.

$X_2$ = Racial/ethnic combination of the ego (respondent) and alter (partner), coded in 6 categories to capture within and across group mixing: black-black, black-Hispanic/white, Hispanic-black/white, Hispanic-Hispanic, white-black/Hispanic, white-white.

$X_3$ = Partnership type (0 = main; 1 = casual).

$X_4$ = The combined age of ego and alter in years.

$X_5$ = The concordant diagnosed HIV-positive status of both ego and alter, compared to all other combinations of dyadic HIV status (1 = concordant positive; 0 = all other combinations of dyadic HIV status).

$X_6$ = Residence (1 = Atlanta metropolitan area; 0 = all other areas).

Note that we modeled the partnership duration and combined age of partners quadratically, and we modeled the interaction of partnership duration and partnership type. Terms within the prediction model were selected based on a combination of *a priori* theory and exploratory data analysis. The coefficients for the model, and their lower and upper 95% confidence intervals, are presented in Supplemental Table 5 below. Exponentiating any linear combination of coefficients will yield the yearly rates, which may be converted to weekly rates through division.

**Supplemental Table 5.** Statistical Model of Act Rates in Main and Casual Partnerships

| Model Parameter | Estimate | Lower 95% CI | Upper 95% CI |
| --- | --- | --- | --- |

| | | | |
|---|---|---|---|
| $\beta_0$ (Intercept) | 4.9615 | 4.9208 | 5.002 |
| $\beta_1$ (Duration) | -0.0013 | -0.0013 | -0.0012 |
| $\beta_2$ (*D*uration$^2$) | 6.3197E-07 | 6.0598E-07 | 6.5781E-07 |
| $\beta_3$ (B-H/W Combo) | 0.5196 | 0.4888 | 0.5505 |
| $\beta_3$ (H-B/W Combo) | 0.2178 | 0.1908 | 0.2449 |
| $\beta_3$ (H-H Combo) | 0.1967 | 0.1687 | 0.2250 |
| $\beta_3$ (W-B/H Combo) | 0.4758 | 0.4505 | 0.5013 |
| $\beta_3$ (W-W Combo) | 0.1765 | 0.1516 | 0.2016 |
| $\beta_4$ (Casual Type) | -1.0373 | -1.0458 | -1.0287 |
| $\beta_5$ (Duration x Casual Type) | -0.0009 | -0.0010 | -0.0009 |
| $\beta_6$ (Combined Age) | -0.0113 | -0.0122 | -0.0104 |
| $\beta_7$ (Combined Age$^2$) | 5.6269E-05 | 5.0154E-05 | 6.2374E-05 |
| $\beta_8$ (HIV+ Concordant) | 0.3614 | 0.3452 | 0.3776 |
| $\beta_9$ (Atlanta residence) | -0.0229 | -0.0396 | -0.0063 |

Abbreviations: CI, confidence interval; B-H/W, black ego with either a Hispanic or white alter; H-B/W, Hispanic ego with either a black or white alter; H-H, Hispanic ego with a Hispanic alter; W-B/H, white ego with either a black or Hispanic alter; W-W, white ego with a white alter.

###### *4.1.3 Predicted Rates in Epidemic Model*

Predicted weekly rates of anal intercourse based on the combination of partnership and individual attributes is then obtained dynamically by predicting from the statistical model with inputs based on the current simulated population. *EpiModel* tracks the current age of partners, the duration of their partnership, their racial combination, and the partnership type. This set of predictors was input into a predict function in R to obtain the weekly mean rates in each stratum. The size of the potential set of strata and corresponding predicted means is therefore nearly infinite based on all the potential combinations of input values.

In Supplemental Figure 1 below, we display some example weekly rates based on a subset of model inputs. This figure shows that rates decline in partnerships with a longer duration, that they are higher in partnerships in which both partners are younger, they are lower for casual partnerships (ptype = 2) compared to main partnerships, and that they are higher in white-white partnerships compared to black-black partnerships. The act rates generally ranged from 0.5 acts per week to 2 acts per week. Other predicted rates may be obtained by exponentiating the coefficients in the table above and dividing by 52 (to convert from yearly rates to weekly rates).

**Supplemental Figure 1.** Predicted Weekly Act Rates from the Poisson Statistical Model, by Partnership Duration, Partnership Type (ptype: 1 = Main; 2 = Casual), Combined Partner Age (comb.age: 40 and 80 Years).

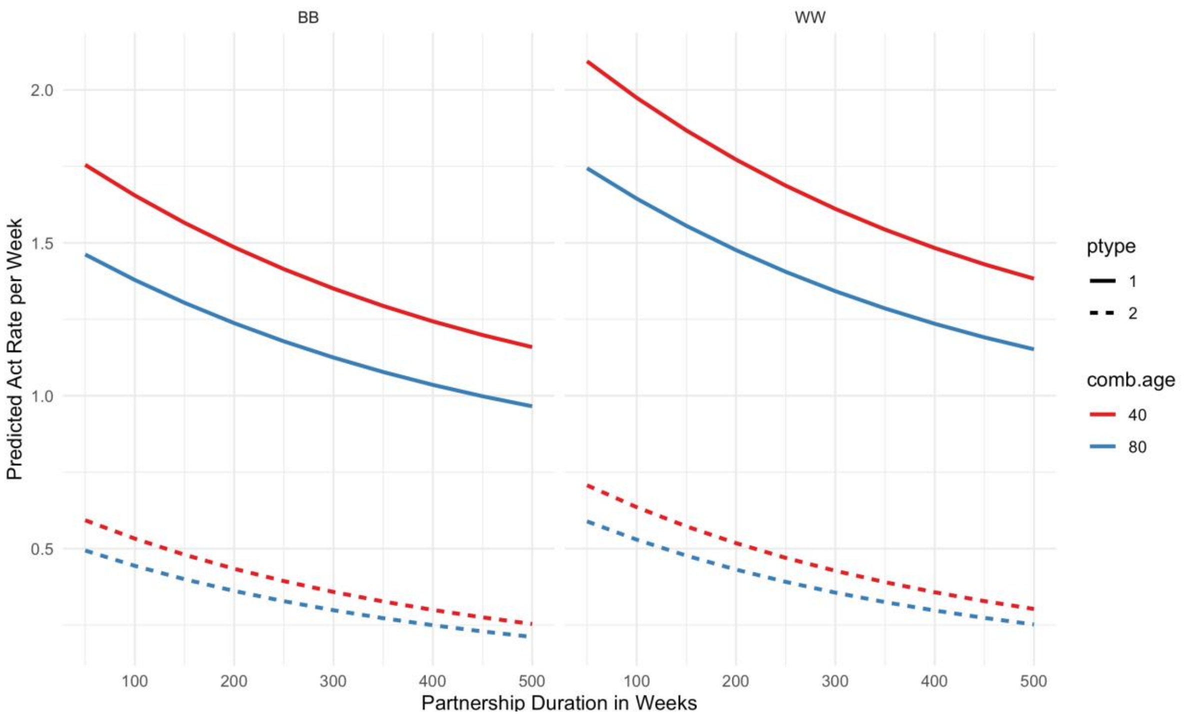

Based on these model predictions, which represent means for each linear combination, we then drew individual counts of acts per partnership per time step in *EpiModel* using the rpois function to draw randomly from the Poisson distribution with a vector of parameters, one value for each partnership.

###### *4.1.4 Cessation of Sexual Activity During Late-Stage AIDS*

In addition to these data-driven statistical calculations, we assumed that MSM in late stages of AIDS (HIV viral load above 5.75), had no acts due to active disease that would limit their sexual activity. This reflected the mid-point between set-point viral load of chronic stage infection (4.5 $\log_{10}$) and peak viral load (7.0 $\log_{10}$, corresponding to the nadir of immunological function). We had no primary data in ARTnet on sexual partnerships in this late disease stage, but prior analysis and modeling studies support a large decline in sexual activity due to AIDS.[17]

##### 4.2 Condom Use Per Act

We modeled condom use within all three partnership types (main, casual, and one-time contacts) based on ARTnet data on the frequency of condom use within reported partnerships. We followed the same general approach to measuring, fitting statistical models, and dynamically predicting condom use within *EpiModel* as we used for rates of anal intercourse. The modeling of condom use here is based on the expectation that changes in condom use depend on race/ethnicity, age, diagnosed HIV status, current PrEP use, and partnership type.

###### *4.2.1 Measurement of Condom Use in ARTnet*

We measured condom use within partnerships in the ARTnet study by asking about the frequency of condom use (for persistent partnerships) or whether condom use occurred (for one-time partnerships) during anal intercourse. Study participants first reported on the number of anal intercourse acts that occurred in the time intervals described above, and then we followed-up with a question on the number of those total acts that involved condom use. We then transformed these subsetted counts into proportions of acts that were condom-protected. This resulted in a U-shaped distribution of proportions, with most persistent partnerships involving either always or never condom use. For this current study, we simplified the outcome variable to any condom use (yes, no) over the past year.

##### *4.2.2 Statistical Models of Condom Use Probabilities*

With the outcome described above, we used the partnership-level dataset to fit two logistic regression models for any condom use in the partnership, with one model for persistent (main and casual) and another model for one-time partnerships. The linear model formula for persistent partnerships was as follows:

$$Y_i \sim \beta_0 + \beta_1 X_1 + \beta_2 X_1^2 + \beta_3 X_2 + \beta_4 X_3 + \beta_5 X_1 X_3 + \beta_6 X_4 + \beta_7 X_4^2 + \beta_8 X_5 + \beta_9 X_6 + \beta_{10} X_7$$

where:

$Y_i$ = Log odds of the probability of condom use per act.

$X_1$ = Duration of partnership in weeks at the survey date.

$X_2$ = Racial/ethnic combination of the ego (respondent) and alter (partner), coded in 6 categories to capture within and across group mixing: black-black, black-Hispanic/white, Hispanic-black/white, Hispanic-Hispanic, white-black/Hispanic, white-white.

$X_3$ = Partnership type (0 = main; 1 = casual).

$X_4$ = The combined age of ego and alter in years.

$X_5$ = The concordant diagnosed HIV-positive status of both ego and alter, compared to all other combinations of dyadic HIV status (1 = concordant positive; 0 = all other combinations of dyadic HIV status).

$X_6$ = Current use of pre-exposure prophylaxis (PrEP) by the ego (respondent) (1 = yes; 0 = no).

$X_7$ = Residence (1 = Atlanta metropolitan area; 0 = all other areas).

Note that we modeled the partnership duration and combined age of partners quadratically, and we modeled the interaction of partnership duration and partnership type. Terms within the prediction model were selected based on a combination of *a priori* theory and exploratory data analysis. The coefficients for the model, and their lower and upper 95% confidence intervals, are presented in Supplemental Table 6 below. Taking the inverse logit of the linear combination of coefficients will yield to the strata-specific predicted probabilities of condom use within the partnership.

**Supplemental Table 6.** Statistical Model of Per Act Condom Use Probability for Main and Casual Partnerships

| **Model Parameter** | **Estimate** | **Lower 95% CI** | **Upper 95% CI** |
|---|---|---|---|
| $\beta_0$ (Intercept) | 2.008 | 1.3020 | 2.7144 |
| $\beta_1$ (Duration) | -0.0031 | -0.0040 | -0.0023 |
| $\beta_2$ ($Duration^2$) | 1.2561E-06 | 5.8878E-07 | 1.8614E-06 |
| $\beta_3$ (B-H/W Combo) | -0.3355 | -0.8549 | 0.1802 |
| $\beta_3$ (H-B/W Combo) | -0.3692 | -0.7798 | 0.04214 |
| $\beta_3$ (H-H Combo) | -0.3989 | -0.8314 | 0.0336 |
| $\beta_3$ (W-B/H Combo) | -0.4402 | -0.8235 | -0.0557 |
| $\beta_3$ (W-W Combo) | -0.5031 | -0.8738 | -0.1310 |
| $\beta_4$ (Casual Type) | 0.5710 | 0.4084 | 0.7347 |
| $\beta_5$ (Duration x Casual Type) | -0.0467 | -0.0638 | -0.0294 |

| | | | |
|---|---|---|---|
| $\beta_6$ (Combined Age) | 0.0002 | 9.5502E-05 | 0.0003 |
| $\beta_7$ (Combined Age$^2$) | -1.6150 | -2.1624 | -1.1322 |
| $\beta_8$ (HIV+ Concordant) | -0.5248 | -0.6790 | -0.3724 |
| $\beta_9$ (PrEP Use) | 0.1701 | -0.1385 | 0.4743 |
| $\beta_{10}$ (Atlanta residence) | 0.0012 | 0.0005 | 0.0019 |

Abbreviations: CI, confidence interval; B-H/W, black ego with either a Hispanic or white alter; H-B/W, Hispanic ego with either a black or white alter; H-H, Hispanic ego with a Hispanic alter; W-B/H, white ego with either a black or Hispanic alter; W-W, white ego with a white alter; PrEP, preexposure prophylaxis.

For the logistic regression model of one-time partnerships, we used a similar logistic regression approach as for persistent partnerships but dropped the partnership duration and partnership type (since there was only one type for this model) predictor variables. The corresponding linear model formula for persistent partnerships was as follows:

$$Y_i \sim \beta_0 + \beta_1 X_1 + \beta_2 X_2 + \beta_3 X_2^2 + \beta_4 X_3 + \beta_5 X_4 + \beta_6 X_5$$

where:

$Y_i$ = Log odds of the probability of condom use per one-time contact.

$X_1$ = Racial/ethnic combination of the ego (respondent) and alter (partner), coded in 6 categories to capture within and across group mixing: black-black, black-Hispanic/white, Hispanic-black/white, Hispanic-Hispanic, white-black/Hispanic, white-white.

$X_2$ = The combined age of ego and alter in years.

$X_3$ = The concordant diagnosed HIV-positive status of both ego and alter, compared to all other combinations of dyadic HIV status (1 = concordant positive; 0 = all other combinations of dyadic HIV status).

$X_4$ = Current use of pre-exposure prophylaxis (PrEP) by the ego (respondent) (1 = yes; 0 = no)

$X_5$ = Residence (1 = Atlanta metropolitan area; 0 = all other areas).

The coefficients for the model, and their lower and upper 95% confidence intervals, are presented in Supplemental Table 7 below. Taking the inverse logit of the linear combination of coefficients will yield to the strata-specific predicted probabilities of condom use within the partnership.

**Supplemental Table 7.** Statistical Model of Per-Act Condom Use Probability for One-Time Sexual Contacts

| **Model Parameter** | **Estimate** | **Lower 95% CI** | **Upper 95% CI** |
|---|---|---|---|
| $\beta_0$ (Intercept) | 2.4287 | 1.6597 | 3.2007 |
| $\beta_1$ (B-H/W Combo) | 0.1526 | -0.3728 | 0.6785 |
| $\beta_1$ (H-B/W Combo) | -0.1042 | -0.5311 | 0.3221 |
| $\beta_1$ (H-H Combo) | -0.10538 | -0.5617 | 0.3506 |
| $\beta_1$ (W-B/H Combo) | -0.1189 | -0.5205 | 0.2825 |
| $\beta_1$ (W-W Combo) | -0.2507 | -0.6414 | 0.1396 |
| $\beta_2$ (Combined Age) | -0.0542 | -0.0733 | -0.0351 |

| | | | |
|---|---|---|---|
| $\beta_2$ (Combined Age$^2$) | 0.0003 | 0.0001 | 0.0004 |
| $\beta_3$ (HIV+ Concordant) | -1.8369 | -2.6547 | -1.1610 |
| $\beta_4$ (PrEP Use) | -0.7133 | -0.8732 | -0.5553 |
| $\beta_5$ (Atlanta residence) | 0.3102 | 0.0107 | 0.6095 |

Abbreviations: CI, confidence interval; B-H/W, black ego with either a Hispanic or white alter; H-B/W, Hispanic ego with either a black or white alter; H-H, Hispanic ego with a Hispanic alter; W-B/H, white ego with either a black or Hispanic alter; W-W, white ego with a white alter; PrEP, preexposure prophylaxis.

##### *4.2.3 Predicted Probabilities in Epidemic Model*

Predicted probabilities of condom use conditional on an anal intercourse act were calculated based on the linear combination of partnership and individual attributes obtained dynamically by predicting from the statistical model

**Supplemental Figure 2.** Predicted Probabilities of Condom Use Per Anal Intercourse Act in Persistent Partnerships from the Logistic Regression Model, by Partnership Duration, Partnership Type (ptype: 1 = Main; 2 = Casual), Combined Partner Age (comb.age: 40 or 80 years), and PrEP Use.

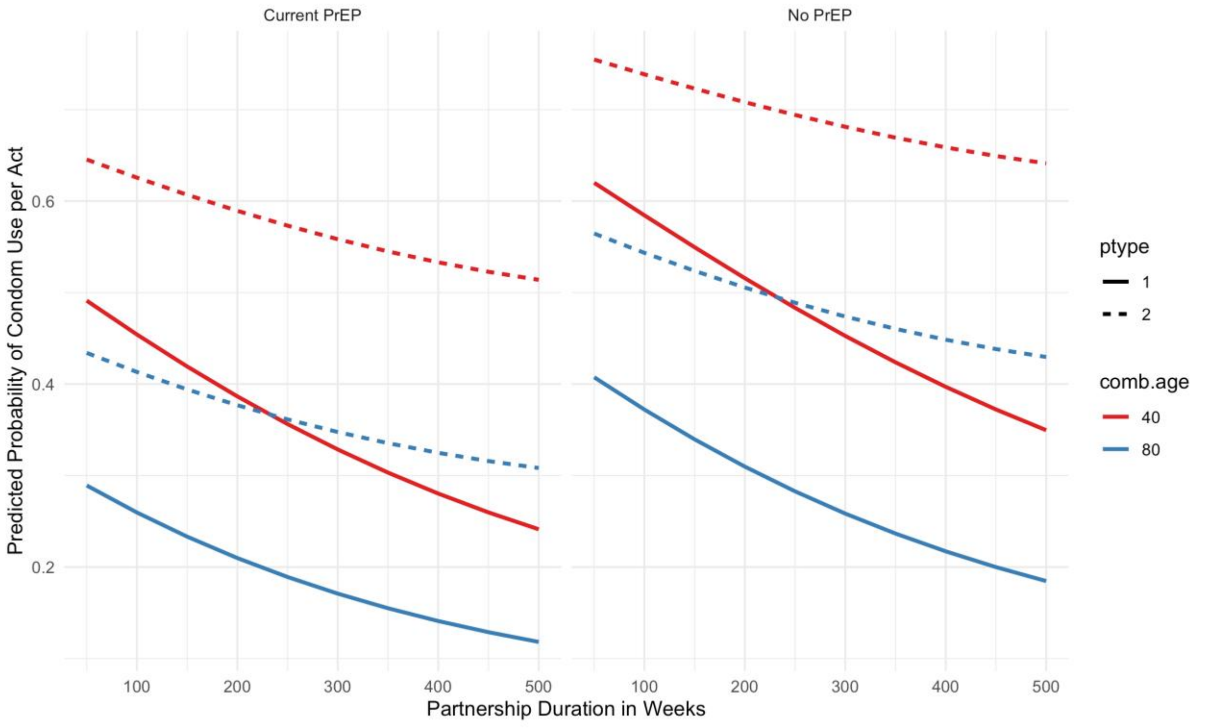

with inputs based on the current simulated population. This set of predictors was input into a predict function in R to obtain the expected mean probabilities. In Supplemental Figure 2, we display some example probabilities based on a subset of model inputs. This figure shows that condom use is lower in partnerships of a longer duration, higher in casual compared to main partnerships, higher when both partners are younger, and lower in partnerships in which the ego (respondent) reported currently using PrEP. Other predicted probabilities may be obtained from Supplemental Table 6 by taking the inverse logit of the linear combination of coefficients of interest.

Supplemental Figure 3 shows the predicted probabilities for the second logistic model, for condom use within one-time anal intercourse contacts. Here we display variation in condom use by combined age of the partners, current PrEP use, and racial combination of the partners. As the figure shows, condom use is higher within partners of a lower combined age, higher in partnerships involving black MSM (race.combo = 1 or 2), and lower among current PrEP users.

Based on these model predictions, which represent expected probabilities for each linear combination, we then drew individual probabilities of condom use per act in *EpiModel* using the rbinom function to draw randomly from the Bernoulli distribution with a vector of parameters, one value for each act. This generated a set of 0's and 1's for whether condom use occurred within the act as a function of the predictors in the statistical model.

**Supplemental Figure 3.** Predicted Probabilities of Condom Use in One-Time Anal Intercourse Contacts from the Logistic Regression Model, by Combined Partner Age, Current PrEP Use, and Racial Combination of Partners (race.combo: 1 = black ego-black alter; 2 = black ego-Hispanic or white alter; 3 = Hispanic ego-black or white alter; 4 = Hispanic ego-Hispanic alter; 5 = white ego-black or Hispanic alter; 6 = white ego-white alter.

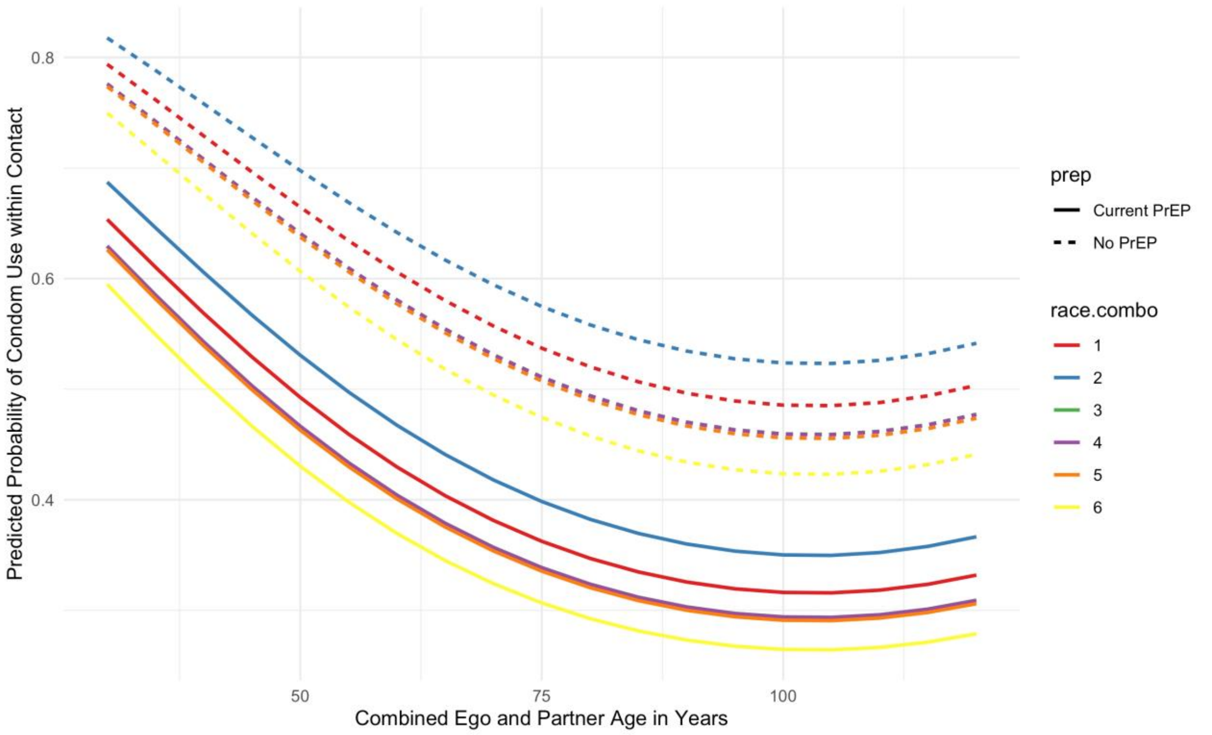

#### 4.4 Sexual Role

Men were assigned an individual sexual role preference (exclusively insertive, exclusively receptive, or versatile) as described in Section 3.1.9. Relationships between two exclusively insertive or two exclusively receptive men are prohibited via the TERGM models. Versatile men were further assigned a preference for being the insertive partner drawn from a uniform distribution between 0 and 1 upon entry into the population; we refer to this proportion as the "insertivity quotient." When two versatile men are simulated to have an anal intercourse act, their sexual positions during that act must be determined (all other allowed combinations have only one direction). One option is for men to engage in intra-event versatility (IEV; i.e. both men engage in insertive and receptive anal intercourse during the act). The probability of this was derived from the partner-specific role data described in Section 3.1.9. If IEV does not occur, then each man's probability of being the insertive partner equals his insertivity quotient divided by the sum of the two men's insertivity quotients.

### 5 DEMOGRAPHY AND INITIAL CONDITIONS

In this model, there are three demographic processes: entries, exits, and aging. Entries and exits are conceptualized as flows into and out of the sexually active population of interest: MSM aged 15 to 65 years old. Entry into this population represents the time at which persons become at risk of infection via male-to-male sexual intercourse, and we model these flows as starting at an age associated with sexual debut and ending at an age potentially before death (age 65). This age range also mapped directly on to the eligibility criteria of the ARTnet study.[8]

#### 5.1 Arrivals at Sexual Onset

All persons enter the network at age 15, which was the lower age boundary of ARTnet. The number of new entries at each time step was based on a fixed rate (0.052 per 100 person-weeks) that kept the overall network size in a relatively stable state. The model parameter governing this rate was tuned iteratively in order to generate simulations with a population size at equilibrium, given the inherent variability in population flows related to background mortality, sexual cessation (i.e., reaching the upper age limit of 65), and disease-induced mortality. At each time step, the exact number of men entering the population was simulated by drawing from a Poisson distribution with the rate parameter.

#### 5.2 Initialization of Attributes

Persons entering the population were assigned attributes in different categories. Some attributes remained fixed by definition (e.g., race/ethnicity), others were fixed by assumption (e.g., insertive versus receptive sexual role), and others were allowed to vary over time (e.g., age and disease status). Here we describe attributes initialized at the outset in the model and for arrivals into the population at each time step:

- **Race/ethnicity.** This model was based on a race/ethnic population composition categorized into three mutually exclusive groups: black, Hispanic, and white. At the outset of the model simulations, individuals were randomly assigned into one of these three groups with a probability equal to the proportions each represented in the Atlanta metropolitan target population based on 2018 Census data estimates for men aged 15 to 65. Those probabilities were: 51.5% black, 4.6% Hispanic, and 43.9% white. Incoming nodes during the dynamic simulation were also randomly assigned a race/ethnicity in these proportions.
- **Age.** In the dynamic simulation, as noted above, all incoming nodes were assigned an age of 15, which incrementally grew in weekly time steps. At the outset of the model simulations, we assigned nodes an age based on a uniform distribution, with ages from 15 to 65. This population-level age distribution was expected to converge to a more realistic distribution during model burn-in and calibration (explained in Section 9.2).
- **HIV Status.** In the dynamic simulation, all incoming nodes were assigned an HIV status of uninfected upon arrival into the population. This reflects the assumption that arrival corresponded with sexual debut, before which exposure to HIV would be very rare. At the outset of the model simulations, we randomly seeded the nodes with HIV infection by fitting and predicting from a logistic regression of diagnosed HIV status from the ARTnet data. This model incorporated city (residence in Atlanta), age, and race/ethnicity as the primary predictors based on the self-reported diagnosed HIV status reported by ARTnet respondents. These initial infections were all assumed to be diagnosed based on this outcome. We did not expect that this initial condition of diagnosed HIV prevalence at the outset of the burn-in model to match the calibrated disease prevalence prior to experimental intervention models; instead this statistical modeling approach allowed for a data-driven seeding of HIV infection in the population that was distributed according to known demographic and geographic heterogeneity. Further description of the transition from initial HIV conditions to calibrated levels are provided in Section 8.2.
- **Circumcision Status.** Circumcision status was randomly assigned to incoming nodes at arrival and for all nodes as initial conditions in the simulations. Based on empirical data from Atlanta MSM,[18] 89.6% of men were circumcised before sexual onset. As described in Section 8, circumcision was associated with a 60% reduction in the per-act probability of infection for HIV-negative males for insertive anal intercourse only (i.e., circumcision did not lower the *transmission* probability if the HIV-positive partner was insertive).[2,19]

#### 5.3 Departures from the Network

All persons exited the network by age 65, either from mortality or by reaching the upper age bound of the MSM target population of interest. This upper limit of 65 was modeled deterministically (probability = 1), but other exits due to mortality were modeled stochastically. Departures included both natural (non-HIV) and disease-induced mortality causes before age 65. Background mortality rates were based on US all-cause mortality rates specific to age and race/ethnicity from the National Vital Statistics life tables.[20] Note that these rates include deaths due to HIV/AIDS; however, the relative fraction of those deaths to total deaths is small enough not to impact this background mortality process. Supplemental Table 8 shows the probability of mortality per year by age and race/ethnicity.

**Supplemental Table 8.** Age- and Race/Ethnicity-Specific Probabilities of Mortality among Men in the United States

| Age | Black | Hispanic | White |
|---|---|---|---|
| 15–19 | 0.00124 | 0.00062 | 0.00064 |
| 20–24 | 0.00213 | 0.00114 | 0.00128 |
| 25–29 | 0.00252 | 0.00127 | 0.00166 |
| 30–34 | 0.00286 | 0.00132 | 0.00199 |
| 35–39 | 0.00349 | 0.00154 | 0.00226 |
| 40–44 | 0.00422 | 0.00186 | 0.00272 |
| 45–49 | 0.00578 | 0.00271 | 0.00382 |
| 50–54 | 0.00870 | 0.00440 | 0.00591 |
| 55–59 | 0.01366 | 0.00643 | 0.00889 |
| 60–64 | 0.02052 | 0.00980 | 0.01266 |

These yearly probabilities were transformed into weekly risks. Natural mortality was then applied to persons within the population at each time step stochastically by drawing from a Bernoulli distribution for each eligible person with a probability parameter corresponding to their age- and race-specific risk of death. Disease-related mortality, in contrast, was modeled based on clinical disease progression, as described in Section 6.

##### 5.4 Aging

The aging process in the population was linear by time step for all persons. The unit of time step in these simulations was one week, and therefore, persons were aged in weekly steps between the minimum and maximum ages allowed (15 and 65 years old). Evolving age impacted background mortality, age-based mixing in forming new partnerships, and other features of the epidemic model described below. Persons who exited the network were no longer active and their attributes such as age were no longer updated.

#### 6 INTRAHOST EPIDEMIOLOGY

Intrahost epidemiology includes features related to the natural disease progression within HIV-positive persons in the absence of clinical intervention. The main component of progression that was explicitly modeled for this study was HIV viral load. In contrast to other modeling studies that model both CD4 and viral load, our study used viral load progression to control both interhost epidemiology (HIV transmission rates) and disease progression eventually leading to mortality.

Following prior approaches,[1,2,4,6,21] we modeled changes in HIV viral load to account for the heightened viremia during acute-stage infection, viral set point during the long chronic stage of infection, and subsequent rise of viral load at clinical AIDS towards disease-related mortality. The HIV viral load has a direct impact on the rates of HIV transmission within serodiscordant pairs in the model, and this interaction is detailed in Section 8. A starting viral load of 0 is assigned to all persons upon infection. From there, the natural viral load curve is fit with the parameters described in Supplemental Table 9.

**Supplemental Table 9.** HIV Natural History Parameters

| Parameter | Value | Reference |
|---|---|---|

| | | |
|---|---|---|
| Time to peak viremia in acute stage | 45 days | Little[22] |
| Level of peak viremia | 6.886 $log_{10}$ | Little[22] |
| Time from peak viremia to viral set point | 45 days | Little,[22] Leynaert[23] |
| Level of viral set point | 4.5 $log_{10}$ | Little[22] |
| Duration of chronic stage infection (no ART) | 3550 days | Buchbinder,[24] Katz[25] |
| Duration of AIDS stage | 728 days | Buchbinder[24] |
| Peak viral load during AIDS | 7 $log_{10}$ | Estimated from average duration of AIDS |

After infection, it takes 45 days to reach peak viremia, at a level of 6.886 $log_{10}$. From peak viremia, it takes another 45 days to reach viral set point, which is set at a level of 4.5 $log_{10}$. Changes occur linearly on the log scale. The total time of acute stage infection is therefore 3 months. The duration of chronic stage infection in the absence of clinical intervention is 3550 days, or 9.7 years. The total duration of pre-AIDS disease from infection is therefore approximately 10 years. At onset of AIDS, HIV viral load rises linearly on the log scale from 4.5 $log_{10}$ to 7 $log_{10}$. The time spent in the AIDS stage is 728 days, or 2 years. This viral load trajectory is for ART-naïve persons only, and the influence of ART on disease progression is detailed in Section 7. These transitions are deterministic for all ART-naïve persons. In the AIDS stage, disease-related mortality is imposed stochastically with a homogenous risk of 1/104, corresponding to average duration of the AIDS stage in weeks. This is accomplished by drawing from a binomial (Bernoulli) distribution for all eligible individuals in the AIDS stage.

#### 7 CLINICAL EPIDEMIOLOGY

Clinical epidemiological processes in the model refer to all steps along the HIV care continuum after initial HIV infection: diagnosis, linkage to ART care, adherence to ART, and HIV viral load suppression. In this model, these clinical features have interactions with the behavioral features detailed above, as well as impacts on the rates of HIV transmission, detailed in the next section. The features of our model's clinical processes generally follow the steps of the HIV care continuum, in which persons transition across states from infection to diagnosis to ART initiation to HIV viral suppression.[26]

##### 7.1 HIV Diagnostic Screening

Both HIV-uninfected and HIV-infected persons in our model were exposed to regular interval-based HIV screening that served as a common entry point for HIV prevention and HIV treatment services, respectively. Individuals screened at routine intervals first based on whether they were currently using PrEP or not. For HIV screening outside of PrEP care, based on exploratory analyses of behavioral and clinical data, and the research questions of this study, we elected to stratify these screening rates by race/ethnicity.

Our approach to parameterization for HIV screening among PrEP non-users was first to start with priors based on ARTnet data for time since last HIV test for HIV-uninfected, and then use model calibration (the technical details of which are explained in Section 9) to fit these parameters to reproduce the race-stratified levels of the first step of the HIV care continuum (the fraction of HIV-infected persons who were diagnosed). For this and the following surveillance target statistics, we have used values specific to MSM. We used this approach because self-reported HIV screening data alone may be biased, and this calibration approach allows for triangulation of diagnostic history based on more objective laboratory data.

Supplemental Figure 4 shows the general results of this calibration. The model starts with all persons with HIV infection as undiagnosed, then the model is simulated for 60 years (x-axis time scale is in weeks) to establish stable equilibrium conditions for this and the other calibrated parameters. The target statistics are shown with dashed horizontal lines and the simulated statistics are shown with solid lines.

**Supplemental Figure 4.** Fraction of MSM with HIV Who Are Diagnosed, Simulations versus Target Statistics, Stratified by Race/Ethnicity (blue = black MSM, red = Hispanic MSM, green = white MSM)

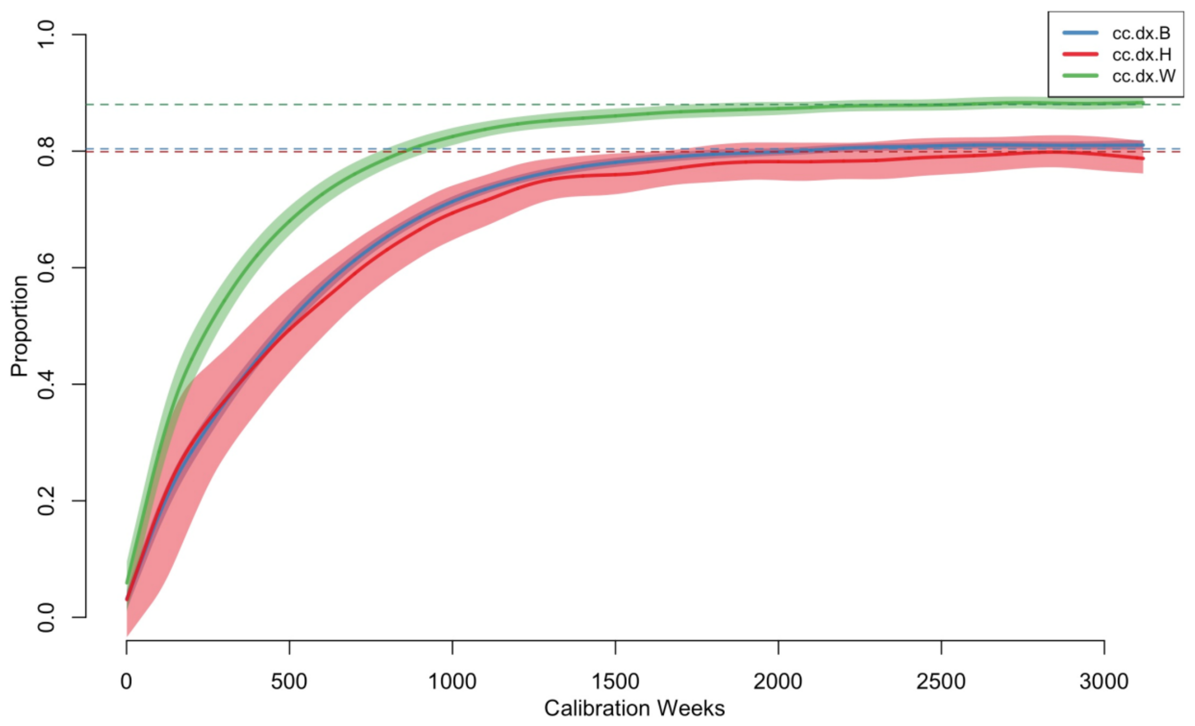

Each model calibration was simulated 1000 times, so the solid lines represent the median values across those simulations and the polygon bands are the interquartile ranges. The three model parameters for the weekly screening rates were calibrated to meet the target statistics, which were the fraction of HIV-infected MSM who were diagnosed. The numerical results from this parameterization are shown in Supplemental Table 10.

**Supplemental Table 10.** Model Parameterization for HIV Screening

| | **Black MSM** | **Hispanic MSM** | **White MSM** |
|---|---|---|---|
| Target Statistic: Diagnosed Fraction[27] | 80.4% | 79.9% | 88.0% |
| Simulations: Diagnosed Fractions | 80.8% | 79.4% | 88.0% |
| Calibrated Rates (per Week) | 0.00385 | 0.00380 | 0.00690 |
| Mean Inter-Test Interval (Years) | 5.00 | 5.06 | 2.79 |
| Median Diagnostic Delay (Years) | 2.50 | 2.52 | 1.70 |

Abbreviation: MSM, men who have sex with men.

The target statistics for the diagnosed fraction were drawn from a Georgia Department of Public Health surveillance report based on laboratory data for MSM in 2017, the most recent year for which the data were available. The diagnosed fraction was higher for white MSM compared to black and Hispanic MSM. After calibration, the simulated diagnosed fractions were nearly identical to those targets. The calibrated screening rates per week were higher among white MSM, and lower among black and Hispanic MSM, consistent with the differentials in the diagnosed fractions across the groups. These weekly rates were consistent with average inter-test intervals, or the average time between HIV negative screening events, of 2.8 to 5.1 years. Note that these intervals represent marginal averages across the target population; some MSM may screen more frequently while others screen very rarely.

We also calculated the diagnostic delay as a validation of this calibration process. Whereas the inter-test interval is calculated for HIV-negative MSM in the model, the diagnostic delay is calculated for HIV-infected MSM who are eventually diagnosed positive. This delay is the median number of years between HIV infection and HIV diagnosis. As shown in Supplemental Figure 5, this time starts out high in the early part of the burn-in model, but converges to a stable equilibrium value by the end of the burn-in. The simulated median values were 2.5 years for black and Hispanic MSM, and 1.7 years for white MSM. This is what would be expected given the differences in the calibrated screening rates. This is also consistent with forward projections of two external studies of national surveillance data. Hall et al. estimate race-stratified median times between infection and diagnosis for 2003 and 2011,[28] and Dailey et al. update these estimates for 2015.[29] The median delays declined substantially over this period, from 5.4 years in 2003 to 3.0 years in 2015. To compare against our other target statistics, we fit a log-linear model to estimate the relative yearly declines in median delay times, with a prediction for 2017. The 2017 projections from this model were 2.44 years overall, 2.47 years for blacks, 2.52 years for Hispanics, and 2.09 years for whites. The corresponding estimates from our simulation model calibrated to the Georgia Department of Public Health HIV care continuum statistics resulted in median times of 2.32 years overall, 2.50 years for blacks, 2.50 years for Hispanics, and 1.70 years for whites. So overall our simulations slightly (by 5%) underestimate the projected 2017 median time to diagnosis, but this gap was small (although larger for white MSM), and it captured the racial/ethnic differences.

**Supplemental Figure 5.** Median Years Delay Between Infection and Diagnosis, Stratified by Race/Ethnicity (blue = black MSM, red = Hispanic MSM, green = white MSM)

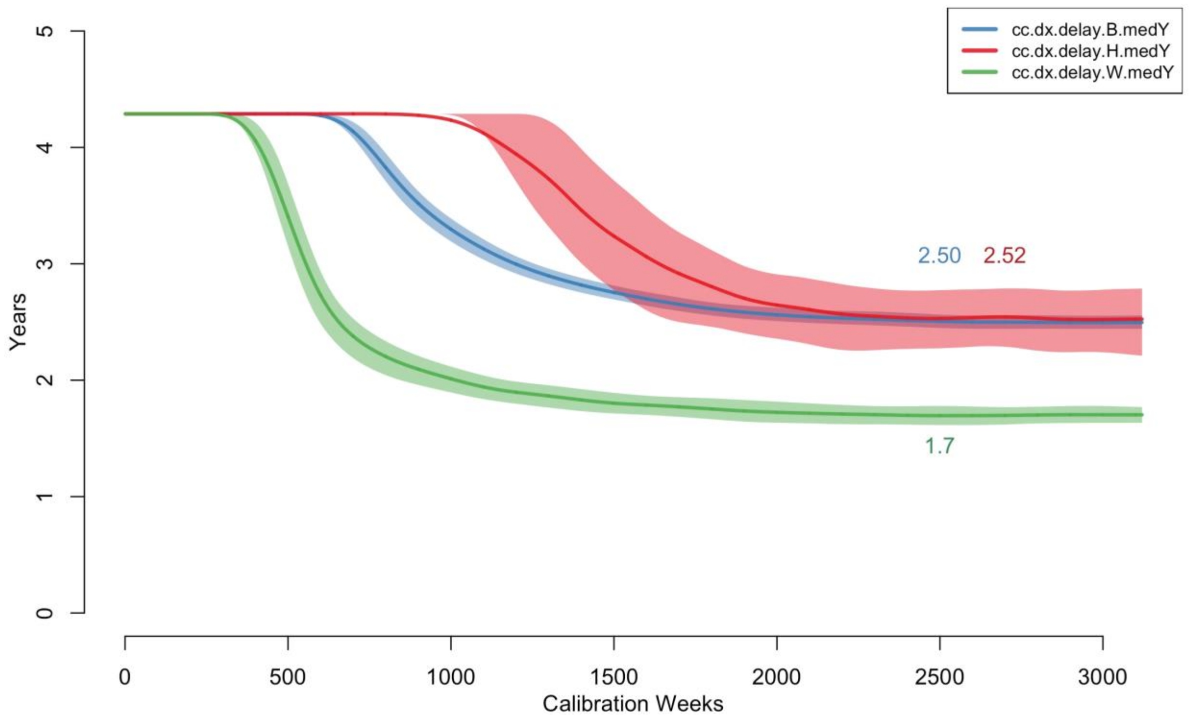

Diagnostic testing was simulated stochastically using draws from a binomial distribution with probability parameters equal to these stratified probabilities. This generated a population-level geometric distribution of times since last test. For PrEP users, we modeled HIV screening practice based on CDC clinical practice guidelines.[30] The guidelines recommend ongoing screening at 3-month intervals for MSM actively using PrEP. This schedule was imposed for all PrEP users active in their PrEP use, regardless of PrEP adherence categories. We also assumed no racial/ethnic variation in HIV screening rates for PrEP users.

Finally, we also modeled a 21-day window period after infection during which the tests of the truly HIV-positive persons would show as negative to account for the lack of antibody response immediately after infection.[31] HIV-positive persons who tested after this window period would be correctly diagnosed with 100% test sensitivity. MSM with recent but undetected infection were still eligible for PrEP initiation since PrEP eligibility was based on diagnosed HIV status. This would have resulted in a period in which HIV-infected but undiagnosed persons were classified as on PrEP. This did not impact their HIV transmission potential (and could not impact their acquisition potential). This undetected infection would then be identified at the next quarterly PrEP clinical visit, at which point they would be transitioned off PrEP.

#### 7.2 Antiretroviral Therapy (ART) Initiation

Following HIV diagnosis, individuals were linked to HIV care that provided ART. In the absence of quantitative data and based on current clinical practice guidelines for MSM in the U.S., we assumed no gap between treatment entry and ART initiation. Although the intermediate steps of the HIV care continuum are often characterized by any linkage to HIV care and/or ART, we selected a second HIV care continuum target of linkage to HIV care specifically within one month of diagnosis for two reasons. First, in the dynamic modeling context, the temporally defined threshold easily mapped on to the tracking implemented for simulated individuals in the model. Second, there were readily available surveillance estimates for this outcome. With respect to the latter, we used data from the Georgia Department of Public Health care continuum estimates for 2017, stratified by transmission risk level and race/ethnicity. We assume therefore that there is a statistical relationship between the proportion linked to care within one month and the average time to care entry following diagnosis: time-to-care entry is assumed to be exponentially distributed, where we use the data on proportion linked to care within one month to solve for the exponential rate parameter. This time-to-event estimate below is generally consistent with recent cohort data that suggest relatively rapid ART initiation following diagnosis.[32]

Supplemental Figure 6 shows the general results to this calibration. The approach was similar to calibration for HIV screening rates. Over the 60-year burn-in simulation period, persons were linked to HIV care with ART with initiation rates that were specific to race/ethnicity. The specific metric used within the simulations to compare against the target statistics was the time period between diagnosis and first ART use, which was uniquely tracked for

**Supplemental Figure 6.** Proportion of Diagnosed HIV-Infected MSM Linked to ART Care within One Month of Diagnosis, Stratified by Race/Ethnicity (blue = black MSM, red = Hispanic MSM, green = white MSM)

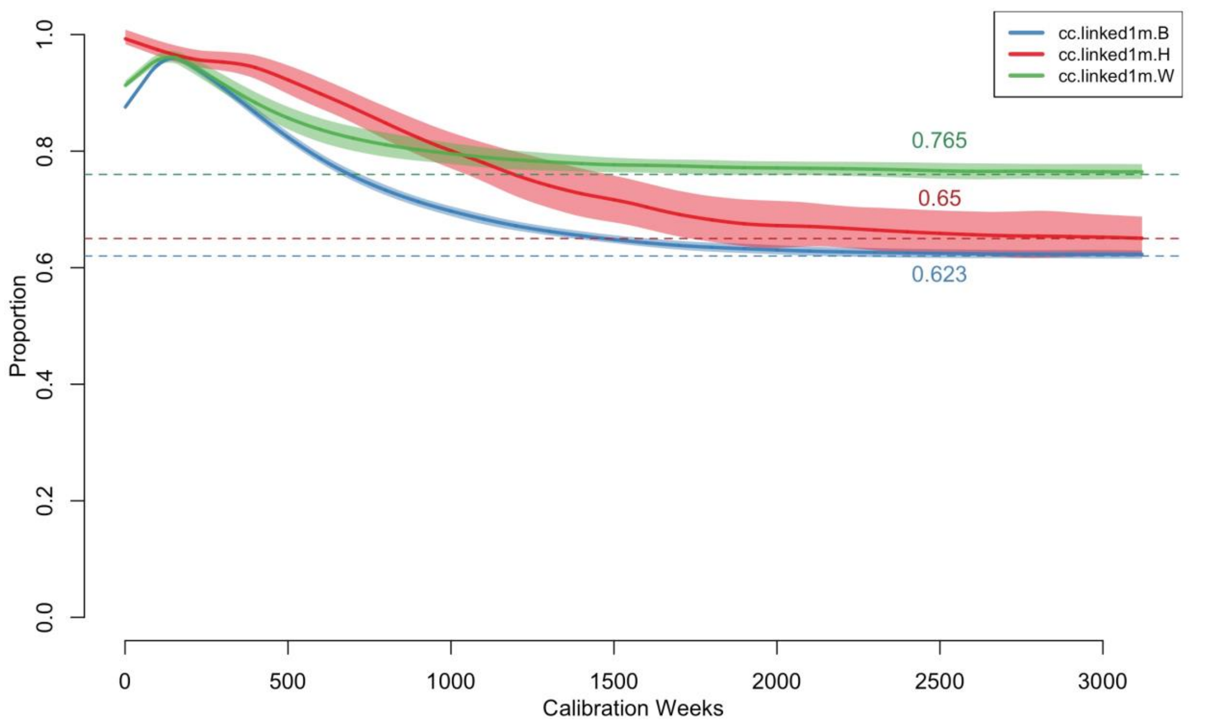

all individuals with HIV infection in the model. A group-specific proportion of persons whose difference between diagnosis and ART initiation was less than or equal to four weeks was calculated in the model. The target statistics are shown with dashed horizontal lines and the simulated statistics are shown with solid lines. Each model calibration was simulated 1000 times, so the solid lines represent the median values across those simulations and the polygon bands are the interquartile ranges.

Supplemental Table 11 shows the numerical results of the calibration. The rate of care establishment was highest for white MSM, and lower for black and Hispanic MSM. With the calibrated rates, the model simulations matched these target statistics. The inverse of these rates implied that the average time to ART initiation after HIV diagnosis was between 4 to 6 weeks on average.

**Supplemental Table 11.** Model Parameterization for ART Linkage After Diagnosis

| | **Black MSM** | **Hispanic MSM** | **White MSM** |
|---|---|---|---|
| Target Statistic: Fraction Linked within 1m[27] | 62% | 65% | 76% |
| Simulations: Fraction Linked | 62.4% | 65.1% | 76.5% |
| Calibrated Rates (per Week) | 0.1775 | 0.1900 | 0.2521 |
| Mean Time to ART (in Weeks) | 5.6 | 5.3 | 4.0 |

Abbreviations: ART, antiretroviral therapy; m, month; MSM, men who have sex with men.

##### 7.3 ART Adherence and HIV Viral Load Suppression

MSM who initiated ART could cycle on and off treatment, where cycling off treatment resulted in an increase in the viral back up to the assumed set point of 4.5 $\log_{10}$. The slope of changes to viral load were calculated such that it took a total of 3 months to transition between the set point and the on-treatment viral loads.[33] Individuals on ART could reach full suppression with sustained ART use. The nadir HIV viral load level was assumed to be 1.5 $\log_{10}$ among those at full suppression levels.[33] The latter corresponds to a rounded value (on the $\log_{10}$) scale of an absolute viral load below the standard levels of detection (viral load = 50).[34] Viral load was tracked and updated continuously over time based on the natural history of HIV disease by stage, and current use of ART.

The patterns of ART adherence (cycling on and off ART) leading to full HIV viral suppression were estimated based on an analysis of HIV care patterns among MSM in the United States[35] and model calibration similar to the first two HIV care continuum steps. The rates of cycling off ART after initially starting (the "halting rate") and the rates of cycling back on after a period of stopping (the "reinitiation rate") controlled overall levels of HIV viral suppression. Within the intervention component of the model, improvement to HIV care retention corresponded to reductions in the halting rate by relative amounts compared to the base calibrates rates.

Because of the negative collinearity of the halting and reinitiation rates that would result in non-identifiability issues if both were simultaneously estimated, we elected to keep the reinitiation rates fixed and fit the halting rates. We started with halting and reinitiation rates and their uncertainty intervals based on an earlier model of the HIV care continuum in the U.S.[36] These reinitiation rates were 0.1326 per year, corresponding to an average time spent off ART before reengagement of 7.5 years. With the reinitiation rates fixed there, we then allowed the halting rates to vary by race/ethnicity and fit them to generate simulations matching the race/ethnicity-specific proportions of diagnosed MSM with a suppressed viral load in the cross-section. We did not model a distinct clinical typology of ART users with a lower propensity for ART discontinuation, above and beyond the differences by race/ethnicity, for two reasons. First, the empirical data to support a distinct typology at the population-level are insufficient. Second, the retention interventions currently in the scenarios are designed to shift the overall population averages rather than focus on a subgroup who would be at higher-risk of ART dropout.

Supplemental Figure 7 shows the general results of this calibration. The general approach was the same as for calibration of HIV screening rates and ART linkage rates. The specific metric used within the simulations to compare against the target statistics was the proportion of individuals who had a HIV viral load below the detectable limit of 200 copies/mL. A group-specific proportion of persons was calculated at each time step in the model. The target statistics are shown with dashed horizontal lines and the simulated statistics are shown with solid lines. Each model calibration was simulated 1000 times, so the solid lines represent the median values across those simulations and the polygon bands are the interquartile ranges.

Supplemental Table 12 shows the numerical results of the calibration. Georgia Department of Public Health data for MSM in 2017 were our target statistics for the proportion of diagnosed MSM with a suppressed viral load in the cross-section. This mapped directly onto to our model simulations.

**Supplemental Figure 7.** Proportion of Diagnosed HIV-Infected MSM with HIV Viral Load Suppression, Stratified by Race/Ethnicity (blue = black MSM, red = Hispanic MSM, green = white MSM)

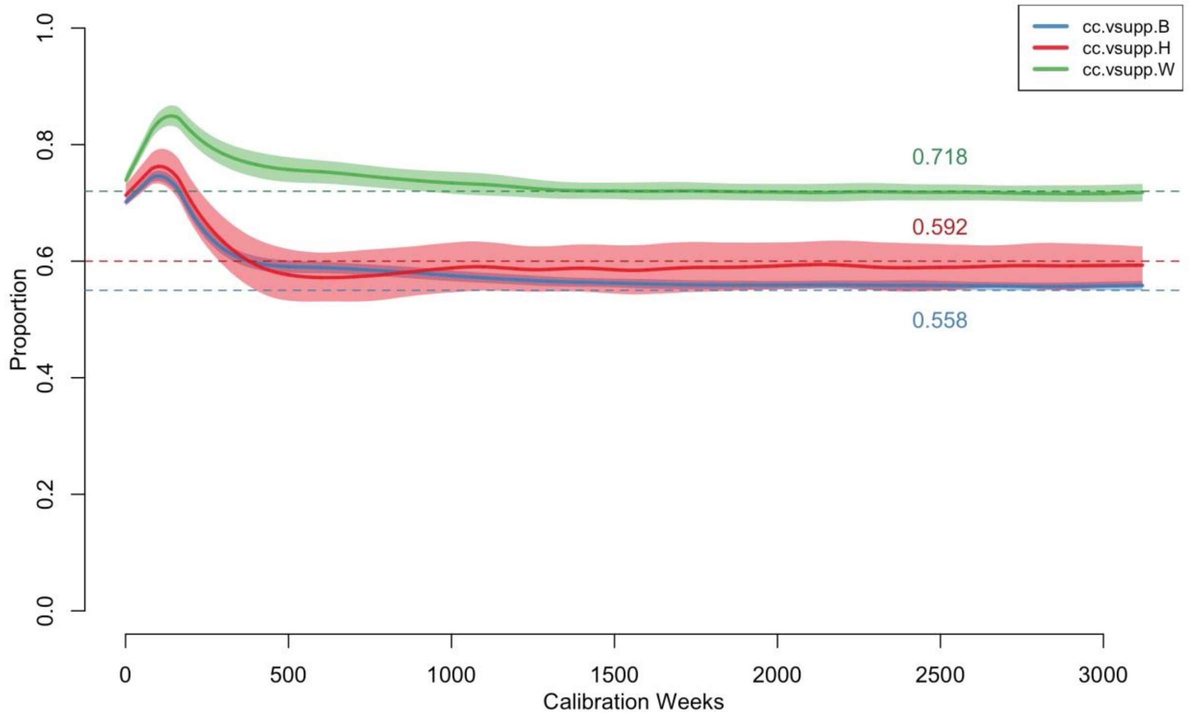

| **Supplemental Table 12.** Model Parameterization for ART Retention Rates After Linkage | | | |
|---|---|---|---|
| | **Black MSM** | **Hispanic MSM** | **White MSM** |
| Target Statistic: Fraction VL Suppressed[27] | 55% | 60% | 72% |
| Simulations: Fraction VL Suppressed | 55.8% | 59.2% | 71.8% |
| Calibrated Halting Rates (per Week) | 0.0062 | 0.0055 | 0.0031 |
| Mean Time to First ART Stoppage (in Weeks) | 161.3 | 181.8 | 322.6 |
| Mean Time to First ART Stoppage (in Years) | 3.1 | 3.5 | 6.2 |

Abbreviations: ART, antiretroviral therapy; MSM, men who have sex with men; VL, viral load.

The corresponding halting rates were therefore lowest in white MSM and highest in black MSM. The inverse of these rates implied a time to first stopping ART after initiation of 161 to 323 weeks.

##### 7.4 AIDS Disease Progression and AIDS-Related Mortality

Progression to AIDS after ART initiation was modeled based on the cumulative time on and off ART for individuals who had been linked to treatment (persons never linked to ART progressed according the rates in Section 6). The maximum untreated time between infection and the start of AIDS for those who never initiate treatment was 9.7 years.[24] For those with some treatment history, we assumed a slower progression time, with individuals who had ever initiated ART spending a maximum of 15 years off of ART over the life course before progression to AIDS, similar to previous models.[1] Persons who had ever initiated ART progressed to AIDS at a similar rate as those who were ART-naïve, but ART use during the AIDS stage was associated with the same declines in HIV viral load as in pre-AIDS stages. However, to account for treatment failure during the AIDS stage, the same mortality rate was applied to persons on active ART and those not on active ART within the AIDS stage. Therefore, we assumed that the probability of disease-induced mortality given AIDS was 1/104 weeks, consistent with approximately 2 years on average spent in the AIDS stage during untreated infection.

#### 8 INTERHOST EPIDEMIOLOGY

Interhost epidemiological processes represent the HIV-1 disease transmission within the model. Disease transmission occurs between sexual partners who are active on a given time step. This section will describe how the overall rate is calculated as a function of the intrahost epidemiological profile of each member of a partnership, and behavioral features within the dyad.

##### 8.1 HIV-Discordant Dyads

At each time step in the simulation, a list of active dyads was selected based on the current composition of the network. This was called an "edgelist." Given the three types of partnerships detailed above, the full edgelist was a concatenation of the type-specific sublists. The complete edgelist reflects the work of the STERGM- and ERGM-based network simulations, wherein partnerships formed on the basis of nodal attributes and degree distributions (see Section 3). From the full edgelist, a disease-discordant subset was created by removing those dyads in which both members were HIV-negative or both were HIV-positive. This left dyads that were discordant with respect to HIV status, which was the set of potential partnerships over which infection may be transmitted at that time step.

##### 8.2 HIV Transmission Rates

Within HIV-discordant dyads, transmission was simulated stochastically across separate sexual acts at each timestep. The per-act probabilities were a combined function of attributes of the HIV-negative and HIV-positive partner. These probabilities were calibrated to reach the empirical diagnosed HIV prevalence. The final per-partnership transmission rates per time step were then a function of one minus these per-act transmission probabilities raised to the number of acts within the partnership during that time step.

###### 8.2.1 Per-Act Transmission Probabilities

Within disease-discordant dyads, HIV transmission was modeled based on a sexual act-by-act basis, in which multiple acts of varying infectiousness could occur within one partnership within a weekly time step. Determination of the number of acts within each discordant dyad for the time step, as well as condom use and role for each of those acts, was described in Section 4. Transmission by act was then modeled as a stochastic process for each discordant sex act following a Bernoulli distribution with a probability parameter that is a multiplicative function of the following predictors of the HIV-negative and HIV-positive partners within the dyad, as shown in Supplemental Table 13 below.

For each act, the overall transmission probability was determined first based on sexual position and HIV viral suppression status of the infected partner. If the infected partner was virally suppressed and on ART, then the base probability was 2.2/100,000, which was derived from a model-based estimate of Supervie.[37] This study estimated the upper bound of the transmission probability of 4.4/100,000 for MSM; we used the mean between the observed number (zero) and this upper bound as our base per-act transmission probability (so 2.2 transmissions per 100,000 exposures) in our model.

If the infected partner was not virally suppressed (at conditions of 200 copies/mL or higher) or not currently on ART, the base probability was a function of whether the HIV-negative partner was in the receptive or insertive role, with the former at a 2.6-fold infection risk compared to the latter. Then, following the parametric function of Wilson,[38] the HIV-positive partner's viral load modifies this base probability in a non-linear formulation, upwards if the viral load was above the viral load set point during chronic stage infection in the absence of ART, and downwards if it was below the set point.

Following others, we modeled an excess transmission risk in the acute stage of infection above that predicted by the heightened VL during that period.[39] Three covariates could reduce the risk of infection: condom use within the act by either the HIV-negative or HIV-positive partner, circumcision status of the HIV-negative partner (only if the HIV-negative partner was insertive in that act), and PrEP use at the time of the act by the HIV-negative partner.

**Supplemental Table 13.** Per-Act Transmission Probabilities and Modifiers

| Predictor | Partner | Parameters | References |
|---|---|---|---|
| Sexual role (insertive or receptive) | HIV- | *Receptive:* 0.008938 base probability when HIV+ partner has 4.5 $\log_{10}$ viral load | Vittinghoff[40] |
| | | *Insertive:* 0.003379 base probability when HIV+ partner has 4.5 $\log_{10}$ viral load | Vittinghoff[40] |
| HIV viral load (VL) | HIV+ (Not virally suppressed or not on ART) | Multiplier of $2.45^{(VL - 4.5)}$ on sexual-role specific base probabilities above | Wilson[38] |
| | HIV+ (Virally suppressed and on ART) | 0.000022 base probability, regardless of sexual role | Supervie[37] |
| Acute stage | HIV+ | Multiplier of 6 | Leynaert,[23] Bellan[39] |
| Condom use | Both | Multiplier of 0.05 times (1 – 0.25) | Varghese,[41] Weller,[42] Smith[43] |
| Circumcision status | HIV-, insertive | Multiplier of 0.40 | Gray[19] |
| Preexposure Prophylaxis (PrEP) | HIV- | High adherence: Multiplier of 0.01<br>Medium adherence: Multiplier of 0.19<br>Low adherence: Multiplier of 0.69 | Grant[44] |

For condom use, we updated our previous approach to explicitly represent condom failure that would result in a transmission event. Our previous models used estimates of HIV incidence comparing consistent condom users to occasional or non-condom users, resulting in a condom "efficacy" of 75–80%. However, this efficacy gap of 20–25% is the function of both the biological/physiological gaps in protection given perfect and consistent condom use during anal intercourse as well as the human error resulting in impact use. Such error could represent condom breakage, misapplication, incomplete use during sexual activity, and other related causes.[43] For this model, we assumed a 95% efficacy for the former, and a 25% absolute reduction in that efficacy as a function of condom failure to arrive at the previous range of 71% total effectiveness.

##### 8.2.2 Calibration of Transmission Probabilities

In addition to the calibration of the HIV care continuum parameters described in Section 7, we also calibrated the per-act transmission probabilities so that the diagnosed HIV prevalence was consistent with empirical data on HIV burden in this target population. Our target statistic for this calibration step was diagnosed HIV prevalence by race/ethnicity, which was estimated in Rosenberg.[45] The target statistics of diagnosed HIV prevalence for MSM in the Atlanta area were 33.3% for black MSM, 12.7% for Hispanic MSM, and 8.4% for white MSM. We took this approach to calibration because there are no external data on the baseline estimated HIV incidence by race/ethnicity for our target population of MSM aged 15 to 65 of all race/ethnicities. There is some historical cohort data for younger (18 to 39 years old) black and white MSM in Atlanta;[12] these were used to calibrate our earlier modeling studies.[4] But we are concerned that the cohort members may be higher risk than all demographically similar MSM in Atlanta due to selection biases. This was a main motivation to calibrate the model primarily based on population-level surveillance targets for the care continuum and disease prevalence.

The per-act transmission probabilities defined above were then multiplied by a factor unique to each race/ethnic group. The final factor levels were 2.21 for black MSM, 0.405 for Hispanic MSM, and 0.255 for white MSM. These calibration factors represent the additional sources of potential error in the transmission parameters that would generate the current HIV epidemic. These include co-factors not included in this model, such as untreated sexually transmitted infections.[46] The upweighting of the transmission probabilities for black MSM and down-weighting for white and Hispanic MSM is due to the long-standing finding that race-stratified behavioral and network data do not, by themselves, explain the excess burden of HIV among black MSM.[47,48]

The results of the calibration are visualized in Supplemental Figure 8. The HIV prevalence was initialized based on the statistical model of diagnosed HIV prevalence with ARTnet data, but allowed to change over the 60-year burn-in period to reach the specified target statistics. In the calibrated model, the median diagnosed HIV prevalence during the final year of the calibration period was 33.1% for black MSM, 12.6% for Hispanic MSM, and 8.5% for white MSM.

**Supplemental Figure 8.** Diagnosed HIV Prevalence, Stratified by Race/Ethnicity (blue = black MSM, red = Hispanic MSM, green = white MSM)

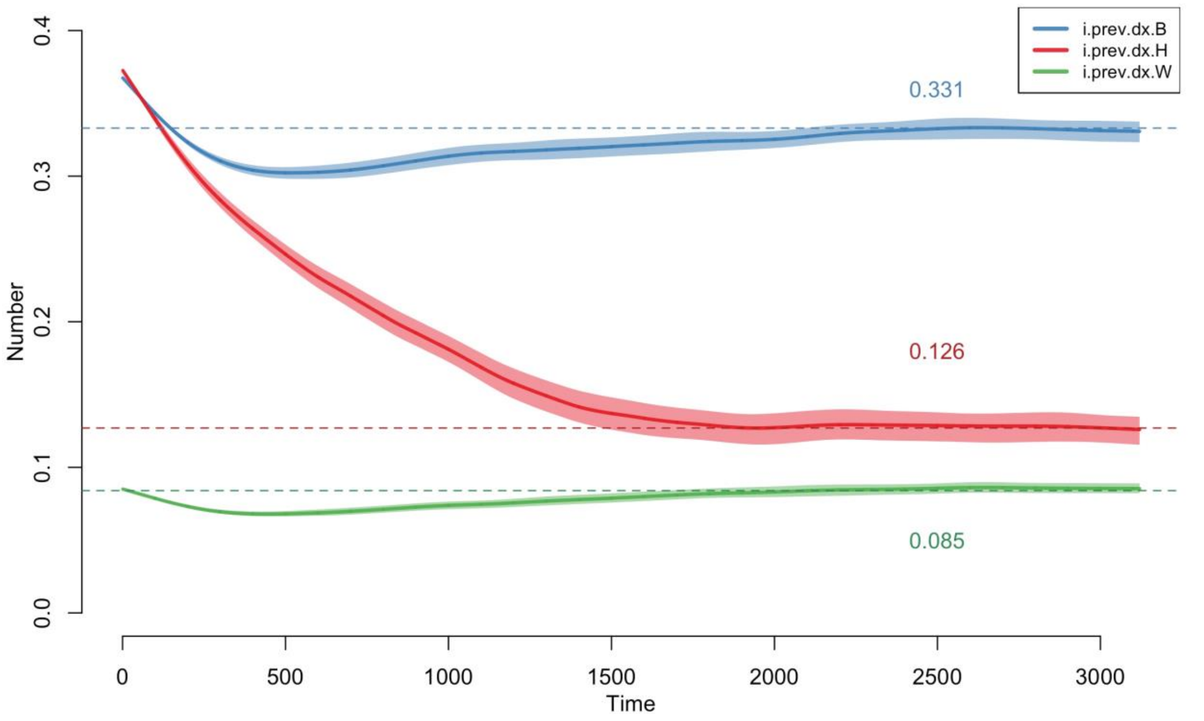

8.2.3 Final Per-Partnership-Week Transmission Rates

The final transmission rate per partnership per weekly time step was a function of the per-act probability of transmission in each act and the number of acts per time step. The per-act transmission probability could be heterogeneous within a partnership due to various types of acts in each interval: for example, a HIV-negative man who is versatile in role may have both insertive and receptive intercourse within a single partnership; some acts within a partnership may be protected by condom use while others are condomless. Transmission was simulated for each act within each serodiscordant dyad, based on draws from a Bernouli distribution with the probability parameter equal to the per-act transmission probabilities detailed above.

#### 9 MODEL CALIBRATION

This section describes the methods for executing the simulations and conducting the data analysis on the outcomes in further detail.

##### 9.1 Calibration Methods

We used Bayesian approaches to define model parameters with uncertain values, construct prior distributions for those parameters, and fit the model to HIV prevalence and incidence data to estimate the posterior distributions of those parameter values.

We used approximate Bayesian computation with sequential Monte Carlo sampling (ABC-SMC) methods[39,49] to calibrate behavioral parameters in which there was measurement uncertainty in order to match the simulated HIV prevalence at the end of the burn-in simulations to the targeted HIV prevalence. The details of ABC depend on the specific algorithm used, but in this case, ABC-SMC proceeded as follows.

For each candidate parameter, $\theta$, to be estimated, we:

1. Sampled a candidate $\theta^i$ from a prior distribution $\pi(\theta)$
2. Simulated the epidemic model with candidate value, $\theta^i$.
3. Tested if a distance statistic, $d$ (e.g., the difference between observed HIV prevalence and model simulated prevalence) was greater than a tolerance threshold, $\epsilon$.
    a. If $d > \epsilon$ then discard
    b. If $d < \epsilon$ then add the candidate $\theta^i$ to the posterior distribution of $\theta$.
4. Sample the next sequential candidate, $\theta^{i+1}$, either independently from $\pi(\theta)$ (if 3a) or from $\theta^i$ plus a perturbation kernel with a weight based on the current posterior distribution (if 3b).

##### 9.2 Calibration Steps

We took a two-step approach to implementing the model calibration. First, we calibrated the model to match the target statistics for the HIV care continuum (screening, linkage, and HIV viral load suppression) and diagnosed HIV prevalence. This involved simulating the model at least 500 times for 60 years (the first burn-in period) and evaluating the distance between the selected target statistics and the simulations at the final year of the period. Once that calibration was complete, we simulated 20,000 replicates of the fitted model and selected the single simulation with the values of the target statistics closest to the targets (with total absolute deviance).

Second, we then simulated the model for an additional 5 years (representing the period between 2013 and 2018) in which PrEP was initially scaled up. The goal of this second burn-in period was to have PrEP coverage (the fraction of eligible MSM who currently use PrEP) calibrated to be approximately 15%. We accomplished this calibration by iteratively adjusting the model parameters for the probability of starting PrEP conditional on eligibility such that the final median PrEP coverage matched this target statistic. For this study, we had to perform this calibration step twice, once for the model scenarios in which it was assumed that PrEP initiation required an HIV-negative screening result, and another for the scenario that assumed that PrEP initiation was random (i.e., it did not require linkage to an HIV-negative screening event). The calibrated probability for the PrEP-linked scenario was 66.0% and the calibrated probability for the PrEP-unlinked scenario was 0.0411%. The probabilities are so different because the opportunities to start PrEP in the unlinked scenario were much higher than in the linked scenario, the latter of which require concurrent indications and an HIV-negative screening event.

#### 10 REFERENCES: SECTIONS 1-9

1 Goodreau SM, Carnegie NB, Vittinghoff E, *et al.* What drives the US and Peruvian HIV epidemics in men who have sex with men (MSM)? *PLoS ONE* 2012; **7**: e50522.

2 Goodreau SM, Carnegie NB, Vittinghoff E, *et al.* Can male circumcision have an impact on the HIV epidemic in men who have sex with men? *PLoS ONE* 2014; **9**: e102960.

3 Carnegie NB, Goodreau SM, Liu A, *et al.* Targeting pre-exposure prophylaxis among men who have sex with men in the United States and Peru: partnership types, contact rates, and sexual role. *J Acquir Immune Defic Syndr* 2015; **69**: 119–25.

4 Jenness SM, Goodreau SM, Rosenberg E, *et al.* Impact of the Centers for Disease Control's HIV preexposure prophylaxis guidelines for men who have sex with men in the United States. *The Journal of infectious diseases* 2016; **214**: 1800–7.

5 Jenness SM, Sharma A, Goodreau SM, *et al.* Individual HIV Risk versus population impact of risk compensation after HIV preexposure prophylaxis initiation among men who have sex with men. *PLoS ONE* 2017; **12**: e0169484.

6 Jenness SM, Weiss KM, Goodreau SM, *et al.* Incidence of gonorrhea and chlamydia following human immunodeficiency virus preexposure prophylaxis among men who have sex with men: A modeling study. *Clin Infect Dis* 2017; **65**: 712–8.

7 Goodreau SM, Hamilton DT, Jenness SM, *et al.* Targeting Human Immunodeficiency Virus Pre-Exposure Prophylaxis to Adolescent Sexual Minority Males in Higher Prevalence Areas of the United States: A Modeling Study. *J Adolesc Health* 2018; **62**: 311–9.

8 Weiss KM, Goodreau SM, Morris M, *et al.* Egocentric Sexual Networks of Men Who Have Sex with Men in the United States: Results from the ARTnet Study. *medRxiv* 2019; : 19010579.

9 Jenness SM, Goodreau SM, Morris M. EpiModel: An R Package for Mathematical Modeling of Infectious Disease over Networks. *J Stat Softw* 2018; **84**: 1–47.

10 Handcock MS, Hunter DR, Butts CT, Goodreau SM, Morris M. statnet: Software Tools for the Representation, Visualization, Analysis and Simulation of Network Data. *J Stat Softw* 2008; **24**: 1548–7660.

11 Zlotorzynska M, Sullivan P, Sanchez T. The Annual American Men's Internet Survey of Behaviors of Men Who Have Sex With Men in the United States: 2016 Key Indicators Report. *JMIR Public Health Surveill* 2019; **5**: e11313.

12 Sullivan PS, Peterson J, Rosenberg ES, *et al.* Understanding racial HIV/STI disparities in black and white men who have sex with men: a multilevel approach. *PLoS One* 2014; **9**: e90514.

13 Krivitsky PN, Handcock MS. A Separable Model for Dynamic Networks. *J R Stat Soc Series B Stat Methodol* 2014; **76**: 29–46.

14 Hunter DR, Handcock MS, Butts CT, Goodreau SM, Morris M. ergm: A Package to Fit, Simulate and Diagnose Exponential-Family Models for Networks. *J Stat Softw* 2008; **24**: nihpa54860.

15 Carnegie NB, Krivitsky PN, Hunter DR, Goodreau SM. An approximation method for improving dynamic network model fitting. *J Comput Graph Stat*; **24**: 502–19.

16 Krivitsky PN, Handcock MS, Morris M. Adjusting for Network Size and Composition Effects in Exponential-Family Random Graph Models. *Stat Methodol* 2011; **8**: 319–39.

17 Hollingsworth TD, Anderson RM, Fraser C. HIV-1 transmission, by stage of infection. *J Inf Dis* 2008; **198**: 687–693.

18 Sullivan PS, Rosenberg ES, Sanchez TH, *et al.* Explaining racial disparities in HIV incidence in black and white men who have sex with men in Atlanta, GA: a prospective observational cohort study. *Ann Epidemiol* 2015; **25**: 445–54.

19 Gray RH, Kigozi G, Serwadda D, *et al.* Male circumcision for HIV prevention in men in Rakai, Uganda: a randomised trial. *Lancet* 2007; **369**: 657–66.

20 United States Census Bureau. Mortality Data. 2012.

21 Jenness SM, Maloney KM, Smith DK, *et al.* Addressing Gaps in HIV Preexposure Prophylaxis Care to Reduce Racial Disparities in HIV Incidence in the United States. *Am J Epidemiol* 2019; **188**: 743–752.

22 Little SJ, McLean AR, Spina CA, Richman DD, Havlir D V. Viral dynamics of acute HIV-1 infection. *J Exp Med* 1999; **190**: 841–50.

23 Leynaert B, Downs AM, de Vincenzi I. Heterosexual transmission of human immunodeficiency virus: variability of infectivity throughout the course of infection. European Study Group on Heterosexual Transmission of HIV. *Am J Epidemiol* 1998; **148**: 88–96.

24 Buchbinder SP, Katz MH, Hessol NA, O'Malley PM, Holmberg SD. Long-term HIV-1 infection without immunologic progression. *AIDS* 1994; **8**: 1123–8.

25 Katz MH, Hessol NA, Buchbinder SP, Hirozawa A, O'Malley P, Holmberg SD. Temporal trends of opportunistic infections and malignancies in homosexual men with AIDS. *J Inf Dis* 1994; **170**: 198–202.

26 Mugavero MJ, Amico KR, Horn T, Thompson MA. The state of engagement in HIV care in the United States: from cascade to continuum to control. *Clin Infect Dis* 2013; **57**: 1164–71.

27 HIV Epidemiology Section, Georgia Department of Public Health. Georgia HIV Care Continuum Update: Persons Living with HIV, and Persons DIagnosed with HIV, 2017. https://dph.georgia.gov/hiv-care-continuum.

28 Hall HI, Song R, Szwarcwald CL, Green T. Brief report: Time from infection with the human immunodeficiency virus to diagnosis, United States. *J Acquir Immune Defic Syndr* 2015; **69**: 248–51.

29 Dailey AF, Hoots BE, Hall HI, *et al.* Vital Signs: Human immunodeficiency virus testing and diagnosis delays—United States. *MMWR Morb Mortal Wkly Rep* 2017; **66**: 1300.

30 Centers for Disease Control and Prevention. Preexposure prophylaxis for the prevention of HIV infection in the United States–2017 Update: A Clinical Practice Guideline. 2017 https://www.cdc.gov/hiv/pdf/risk/prep/cdc-hiv-prep-guidelines-2017.pdf.

31 Fiebig EW, Wright DJ, Rawal BD, *et al.* Dynamics of HIV viremia and antibody seroconversion in plasma donors: implications for diagnosis and staging of primary HIV infection. *AIDS* 2003; **17**: 1871–9.

32 Medland NA, Chow EPF, McMahon JH, Elliott JH, Hoy JF, Fairley CK. Time from HIV diagnosis to commencement of antiretroviral therapy as an indicator to supplement the HIV cascade: Dramatic fall from 2011 to 2015. *PLoS ONE* 2017; **12**: e0177634.

33 Chu H, Gange SJ, Li X, *et al.* The effect of HAART on HIV RNA trajectory among treatment-naive men and women: a segmental Bernoulli/lognormal random effects model with left censoring. *Epidemiology* 2010; **21 Suppl 4**: S25-34.

34 Chun T-W, Carruth L, Finzi D, *et al.* Quantification of latent tissue reservoirs and total body viral load in HIV-1 infection. *Nature* 1997; **387**: 183–8.

35 Beer L, Oster AM, Mattson CL, Skarbinski J. Disparities in HIV transmission risk among HIV-infected black and white men who have sex with men, United States, 2009. *AIDS* 2014; **28**: 105–14.

36 Shah M, Perry A, Risher K, *et al.* Effect of the US National HIV/AIDS Strategy targets for improved HIV care engagement: a modelling study. *Lancet HIV* 2016; **3**: e140-146.

37 Supervie V, Breban R. Brief Report: Per Sex-Act Risk of HIV Transmission Under Antiretroviral Treatment: A Data-Driven Approach. *J Acquir Immune Defic Syndr* 2018; **79**: 440–4.

38 Wilson DP, Law MG, Grulich AE, Cooper DA, Kaldor JM. Relation between HIV viral load and infectiousness: a model-based analysis. *Lancet* 2008; **372**: 314–20.

39 Bellan SE, Dushoff J, Galvani AP, Meyers LA. Reassessment of HIV-1 acute phase infectivity: accounting for heterogeneity and study design with simulated cohorts. *PLoS Med* 2015; **12**: e1001801.

40 Vittinghoff E, Douglas J, Judson F, McKirnan D, MacQueen K, Buchbinder SP. Per-contact risk of human immunodeficiency virus transmission between male sexual partners. *Am J Epidemiol* 1999; **150**: 306–11.

41 Varghese B, Maher JE, Peterman TA, Branson BM, Steketee RW. Reducing the risk of sexual HIV transmission: quantifying the per-act risk for HIV on the basis of choice of partner, sex act, and condom use. *Sex Transm Dis* 2002; **29**: 38–43.

42 Weller S, Davis K. Condom effectiveness in reducing heterosexual HIV transmission. *Cochrane Database Syst Rev* 2002; : CD003255.

43 Smith DDK, Herbst JHJ, Zhang X, Rose CE. Condom effectiveness for HIV prevention by consistency of use among men who have sex with men in the United States. *J Acquir Immune Defic Syndr* 2015; **68**: 337–44.

44 Grant RM, Anderson PL, McMahan V, *et al.* Uptake of pre-exposure prophylaxis, sexual practices, and HIV incidence in men and transgender women who have sex with men: a cohort study. *Lancet Infect Dis* 2014; **14**: 820–9.

45 Rosenberg ES, Purcell DW, Grey JA, Hankin-Wei A, Hall E, Sullivan PS. Rates of prevalent and new HIV diagnoses by race and ethnicity among men who have sex with men, U.S. states, 2013-2014. *Ann Epidemiol* 2018; **28**: 865–73.

46 Bernstein KT, Marcus JL, Nieri G, Philip SS, Klausner JD. Rectal gonorrhea and chlamydia reinfection is associated with increased risk of HIV seroconversion. *J Acquir Immune Defic Syndr* 2010; **53**: 537–43.

47 Millett GA, Peterson JL, Wolitski RJ, Stall R. Greater risk for HIV infection of black men who have sex with men: a critical literature review. *Am J Public Health* 2006; **96**: 1007–19.

48 Goodreau SM, Rosenberg ES, Jenness SM, *et al.* Sources of racial disparities in HIV prevalence in men who have sex with men in Atlanta, GA, USA: a modelling study. *Lancet HIV* 2017; **4**: e311–e320.

49 Toni T, Welch D, Strelkowa N, Ipsen A, Stumpf MPH. Approximate Bayesian computation scheme for parameter inference and model selection in dynamical systems. *J R Soc Interface* 2009; **6**: 187–202.

#### 11 INTERVENTIONS AND ANALYSIS-SPECIFIC DETAILS

Sections 11-13 of this Supplemental Appendix provide details about methods for the cost-effectiveness analysis, as well as additional results from sensitivity analyses.

##### 11.1 Initiation Intervention

###### *11.1.1 Background*

*HealthMindr* is a smartphone application that is currently under evaluation in a clinical trial that aims to increase PrEP uptake among users who are indicated for PrEP (ClinicalTrials.gov Identifier: NCT03763942).[1] Participants are being recruited from Jackson, MS, Atlanta, GA, and Washington DC, are between ages 18-34, and 50% of participants are racial/ethnic minorities.

The smartphone application includes access to HIV testing, non-occupational post-exposure prophylaxis (nPEP), behavioral risk assessments, PrEP, product ordering (products can be ordered directly and anonymously from Amazon), and a substance use directory. Users also complete a monthly PrEP eligibility assessment, and if a user is eligible for PrEP, they will be referred to nearby PrEP providers. The clinical trial also included quarterly surveys to assess demographics and behavior, electronic reminders to complete surveys, option to have at home HIV tests twice/year, option to have at-home STI tests twice/year, alcohol and substance use screening, and tenofovir detection.

We assume that the intervention when implemented outside a clinical trial would include only a subset of services used in the trial, specifically: access to the smartphone application, monthly PrEP eligibility assessments, at-home HIV and STI tests, and product ordering.

###### *11.1.2 Model Implementation*

In our model, MSM who were HIV- or undiagnosed HIV+ and not already using PrEP were eligible to start *HealthMindr*. At each weekly timestep, all eligible individuals experienced a uniform probability of app initiation. It was assumed that *HealthMindr* users engaged with the app for 3 months then returned to the app-eligible population. The weekly probabilities for *HealthMindr* initiation were selected to yield coverage levels of app usage that matched our low (12%), base case (22%), and high (30%) coverage assumptions given the assumed app discontinuation at 3 months. MSM using the *HealthMindr* app had an additional PrEP initiation probability (elevating the baseline probability) that corresponded to the intervention efficacy.

###### *11.1.3 Effectiveness*

The estimation of the weekly PrEP initiation probability experienced by *HealthMindr* app users was derived from assumptions in the sample size calculation of the *HealthMindr* study protocol.[1] The calculations assumed the hazard of PrEP initiation in the intervention arm was 4.5 times higher than that of the control arm. Using the model parameters required to achieve 15% PrEP coverage (the estimated baseline coverage for Atlanta), the non-intervention hazard rate of PrEP initiation was first calculated (0.195/year). This hazard rate was multiplied by 4.5 (0.195*4.5 = 0.8775/year) and converted to a weekly probability to obtain the total probability of PrEP initiation for a *HealthMindr* app user each week (p = 1-exp(-0.8775/52) = 0.01673). This total probability is the result of both the app use and routine HIV testing not related to app use. To separate out the PrEP initiation probability due to the app alone, the annual non-intervention initiation rate was subtracted from the annual total intervention rate (0.8775-0.195= 0.6852/year), and converted to a weekly probability: p=1-exp(-0.6852/52) = 0.013. Therefore, in the simulation, indicated individuals experienced an additional 0.013 probability of initiating PrEP each week when using the app.

##### 11.2 Adherence Intervention

###### *11.2.1 Background*

*Life-Steps for PrEP* was a trial conducted in Boston, MA to improve PrEP adherence among MSM.[2] Trial components included 9 clinic visits (6 of which include counseling) over 6 months, Wisepill real-time adherence monitoring, tenofovir measures, self-interviews on sexual behavior, and daily text messages to assess sex and condom behavior. We assume the intervention as implemented outside a clinical trial would include 6 clinic visits/counseling sessions over three months as well as educational materials.

##### 11.2.2 *Model Implementation*

In our model, MSM were eligible for *Life-Steps* at PrEP initiation. Under the low, base case, and high coverage assumptions, we assumed that 15%, 30%, or 60% of new PrEP users would complete the *Life-Steps* intervention. This intervention had the effect of shifting the probability of being assigned to high PrEP adherence from 60% (baseline) to 87.1% for intervention participants. The overall distribution of adherence levels in the status quo was 16.5% of PrEP users with low adherence, 23.5% medium, and 60% high. With the intervention, adherence levels were 5.3% low, 7.6% medium, and 87.1% high. This was a one-time intervention with effects lasting the duration of an episode of PrEP use. Individuals could participate in this intervention more than once if they had multiple PrEP episodes during the 10-year model intervention period.

##### 11.2.3 *Effectiveness*

The estimation of the proportion of individuals in the high adherence category when engaged in the *Life-Steps* intervention was based on pilot study results:[2] at the two assessment points, on average 87.1% of participants had high adherence (90.0% at 3 months and 84.2% at 6 months) defined as plasma tenofovir levels reflective of perfect PrEP adherence.

#### 11.3 Persistence Intervention

##### 11.3.1 *Background*

*PrEP@Home* and *ePrEP* are interventions focused on improving MSM retention on PrEP by allowing patients to have home-based rather than clinic-based PrEP monitoring and follow-up.[3,4] Similar to *HealthMindr*, *PrEP@Home* and *ePrEP* are still in the trial phase (ClinicalTrials.gov Identifiers: NCT03729570, NCT03569813).

The intervention includes a patient-facing app, where patients can track and confirm test kit receipt, access instructions for taking tests, complete surveys, and receive reminders to take tests and send them back in a timely manner. There is also a clinician dashboard where clinicians can view patient's home-based test results and renew PrEP prescriptions without a clinic visit if all lab values are normal. At home test kits contain tests for creatine levels, HIV, chlamydia, gonorrhea, and syphilis, as well as testing instructions and pre-paid mailers to return the tests. Telemedical consults are provided as needed for participants.

We assume that the intervention when implemented outside a clinical trial would include the patient app, home test kits, the clinician dashboard, and telemedicine visits at PrEP initiation and once per year while patients remain on PrEP.

##### 11.3.2 *Model Implementation*

In our model, MSM were eligible for the persistence intervention at PrEP initiation. Under the low, base case, and high coverage assumptions, we assumed that 20%, 40%, or 80% of newly initiated PrEP users would start the intervention. This intervention had the effect of reducing the weekly probability of random PrEP discontinuation (i.e., not due to a change in indications), such that the duration of an episode of PrEP use was increased. We assumed that enrollment in the persistence intervention was a one-time occurrence and that engagement in the persistence intervention lasted the duration of a PrEP episode. Individuals could participate in this intervention more than once if they had multiple PrEP episodes during the 10-year model intervention period.

##### 11.3.3 *Effectiveness*

The median duration of a PrEP episode and the corresponding weekly probability of discontinuation for those engaged in the persistence intervention were derived using assumptions in the *PrEP@Home* study protocol obtained from trial investigators.[3,4] The minimum clinically relevant treatment effect assumed in the protocol's sample size calculation corresponds to an absolute increase in the proportion of PrEP users retained after 1 year of 15.8%. The baseline weekly probability of discontinuation (0.02139) was used to calculate the non-intervention proportion of PrEP users retained through 1 year (32.5%). The assumed treatment effect of the intervention was added to this proportion (32.5%+15.8%=48.3%), and a new lower weekly probability of discontinuation (0.01389) was calculated from this higher proportion of retention. This lower weekly probability of discontinuation results in a median duration of PrEP of 346.7 days in the model.

##### 11.4 Health State Utilities and QALYs

Health state utilities are based on an individual's HIV infection status, CD4 count, ART status, viral suppression status, and age (Supplemental Table 1).[5] At each weekly time-step in the model simulation, individuals accrued quality-adjusted life-weeks based on these utilities, which were then aggregated into quality-adjusted life-years (QALYs). Age-specific utility was assumed to following a linearly decreasing function, decreasing by 0.003 each year.[5] An individual's age-specific utility reduction was subtracted by their health state-specific utility to calculate their overall utility. For example, a 25-year-old that is uninfected and not on PrEP would have a health state utility of 1.0 (perfect health), which would be reduced by 0.003*25 = 0.075 for an overall utility of 0.925. A 25-year-old that is on ART, virally suppressed, with CD4<200 would have a health state utility of 0.87, for an overall utility of 0.87-0.075 = 0.795. QALYs accrued within the simulation were discounted at an annual rate of 3%.

**Supplemental Table 1.** Health State Utilities

| Individual Health State | Utility Value |
|---|---|
| Uninfected, Not on PrEP | 1 |
| Uninfected, On PrEP | 1 |
| On ART, Virally Suppressed, CD4<200 | 0.87 |
| HIV+, On ART, Virally Suppressed, CD4>200 | 0.89 |
| AIDS, On ART, Virally Suppressed, CD4<200 | 0.85 |
| HIV+, On ART, Not Virally Suppressed, CD4>200 | 0.89 |
| AIDS, Stopped ART, CD4<200 | 0.82 |
| HIV+, Stopped ART, CD4>200 | 0.87 |
| AIDS, Never Started ART, CD4<200 | 0.83 |
| HIV+, Never Started ART, CD4>200 | 0.96 |
| Age-Adjustment | -0.003 |

PrEP: pre-exposure prophylaxis; ART: antiretroviral therapy.

##### 11.5 Analytic Time Horizon

The time horizon is composed of three sections in the model: 1) Intervention period: in the first 10 years, all modules described in Appendix A (disease transmission, sexual behavior, population entry, etc.) are enabled and relevant interventions are active; 2) Post-intervention period: in the next 10 years (years 10-20), all modules are enabled, but the interventions are disabled; this reflects a post-intervention period where interventions are not active but they may still have downstream effects on transmission as PrEP cascade levels eventually return to pre-intervention levels; 3) End-horizon period: at the end of the post-intervention period, anyone alive is simulated until death to capture their remaining lifetime costs and QALYs. During the end-horizon period, disease transmission and sexual behavior modules are disabled and no new individuals enter the model; however, HIV testing, treatment, and disease progression modules are still enabled because these dynamics would persist in the absence of further disease transmission and impact an individual's remaining costs and QALYs.

##### 11.6 Intervention Cost Estimates

Personnel, overhead, direct medical, and technology/material resource costs were estimated based on intervention components and assumptions on the time and resources needed to carry out each component. Each intervention's cost estimates included both fixed costs (same cost regardless of number of users) and per-person variable costs (scales with number of users). As a simplification, per-person cost for the entire duration of a user's engagement of the intervention were assumed to be incurred at initiation (either beginning use of the initiation app or at PrEP initiation).

For all interventions, personnel wages were estimated from US Bureau of Labor Statistics median wage estimates for the Atlanta-Sandy Springs-Roswell region for relevant occupations (Table 2).[6] We assumed that wages reflected 70% of total compensation[7] and therefore multiplied wage rates by 1/0.7 to calculate total personnel costs, which includes employer-provided fringe benefits, such as health insurance and retirement contributions. For example, 6 hours of registered nurse time would be calculated as ($34.82/0.7)*6 = $298. For consistency across the analysis, intervention costs were updated to 2020 USD using the Consumer Price Index for medical care.

| **Supplemental Table 2.** Wages by Occupation in Atlanta-Sandy Springs-Roswell (2018) | | | | | | |
|---|---|---|---|---|---|---|
| **Code** | **Occupation** | **Hourly Wage** | | | | **Mean Annual** |
| | | **25th** | **50th** | **75th** | **Mean** | |
| 29-1141 | Registered Nurses | $29.28 | $34.82 | $40.37 | $35.19 | $73,190 |
| 21-1022 | Healthcare social workers | $21.59 | $27.52 | $33.45 | $27.39 | $56,980 |
| 15-1132 | Software Developers, Applications | $36.90 | $49.13 | $60.26 | $51.18 | $106,450 |
| 29-1062 | Family and General Practitioners | - | $95.54 | - | $104.47 | $217,290 |
| 21-1093 | Social and human service assistants | $10.50 | $13.85 | $17.89 | $14.60 | $30,360 |

Source: US Bureau of Labor Statistics. May 2018 Metropolitan and Nonmetropolitan Area Occupational Employment and Wage Estimates: Atlanta-Sandy Springs-Roswell. Accessed 2/29/2020. GA. https://www.bls.gov/oes/current/oes_12060.htm#29-0000

##### *11.6.1 Initiation intervention*

In the base case, we assumed that the PrEP initiation intervention personnel costs consisted of 10 hours/month of software engineering ($741.16) and 16 hours/month of app maintenance (e.g., ensuring PrEP resources and provider information is up-to-date on the app) performed by a social and human services assistant ($334.30). We assumed that there were no overhead costs associated with the app because there are no location-based services. For direct medical, in the the base case, we assumed that each participant received 2 HIV tests, 2 STI tests, 2 condom orders, and 1 lube order per year when using the app [2($47) + 2($228.82)+2($10)+1($9.25) = $352.07/year or $29.34/month]. Costs of these services and shipping were estimated from study cost data collected by the *HealthMindr* team (Table 3). For material resources, we assume fixed costs associated with server fees ($500/month) and messaging fees ($1,009/year) for the smartphone application using cost data provided by *HealthMindr*. We also assumed per-person costs associated with online advertising to get new users onto the app. Using data from *HealthMindr*, we estimate that it costs $0.79 to get a person to click to learn more about the app, and then we assume 0.1% of people who click on an ad end up as app users, which comes to $84.32 in advertising costs per new app user.

Thus, the initiation intervention had a monthly fixed cost that included software engineering and human services personnel ($741.16 + $334.30 = $1,075.46/month) and messaging/server resource costs ($1,009/12 + $500 = $584.08/month), for a total of $1075.46+$584.08 = $1,659.54/month. A one-time per person cost was also incurred when individuals started using the app, which combined the advertising cost to reach users ($84.32 per person) and direct medical costs, assuming individuals use the app for an average of 3 months ($29.34*3 = $88.02), for a total of $84.32+$88.02= $172.34 per new user.

| **Supplemental Table 3.** Unit Costs for Direct Medical and Resource Components of Interventions | |
|---|---|
| **Item** | **Unit Cost** |
| HIV Test Kit | $40.00 |
| HIV Test Shipping | $7.00 |
| STI Test Kit | $17.85 |
| STI Test Shipping | $16.25 |
| STI Lab Fee | $80.31 |
| Condoms | $2.00 |
| Condoms Shipping | $8.00 |
| Lube | $1.25 |
| Lube Shipping | $8.00 |
| Creatinine Test | $9.46* |
| Creatinine Test Shipping | Assume bundled in HIV shipping |
| App Text Messaging Fees | $1,009/year |
| Server/App Hosting Fees | $500/month |

Source: Preliminary study cost data from *HealthMindr* team sent on March 2, 2020.
*Creatinine test cost is from CMS Clinical Laboratory Fee Schedule 2020.

##### 11.6.2 *Persistence intervention*

In the base case, the PrEP persistence intervention personnel costs consisted of fixed costs of 10 hours/month for software engineering/app support ($741.16), 1 telemedicine appointment at PrEP initiation ($77.17), 10 minutes of physician time per participant every 3 months to review lab results and refill PrEP prescriptions ($28.50), and 1 telemedicine appointment per person per year for PrEP follow-up visits ($52.29). No overhead costs were included because care was provided entirely virutally. For direct medical costs, we assumed 4 at-home HIV tests, 2 at-home STI tests, and 2 at-home creatinine tests per person per year, based on CDC guidelines for PrEP-associated testing [4($47)+2($114.41)+2($9.46) = $435.74/year]. For material resources, we assumed that intervention would incur text messaging service fees and server/hosting fees as fixed costs at the same level as recorded by *HealthMindr* [$1,009+12($500) = $7,009/year or $584/month].

The monthly total fixed cost consists of material resource and software engineering personnel costs [$584+$741 = $1,325/month]. The one-time per person cost assumes users are retained on the intervention for an average of 12 months, and combines personnel [$77.17+4($28.50)+$52.29 = $243.49] and direct medical [$435.74/year] for a total cost of $243+$435.74 = $679.25.

##### 11.6.3 *Adherence intervention*

In the base case, personnel costs associated with the PrEP adherence intervention was based on participants attending six 50-minute nurse-led behavioral counseling sessions, which is costed at 6 visits x 60 minutes of registered nurse time (6*$52.53 = $315.17/person), using median nurse wages. In addition to personnel time, an overhead cost was included based on hospital outpatient reimbursement rate for a clinic visit and related services (APC code 5012, using relevant local labor index value for Atlanta), since the counselling sessions would require the use of private clinic space. We used APC code 5012 for pay rate and the local labor index for Atlanta to estimate an overhead cost of $110.72 per counselling session, for a total of $664.32/person for the full six-session intervention. The adherence intervention incurred no direct medical costs, as in practice, participants do not receive medical care or prescriptions as part of the intervention. For material resources, we assumed that counselling sessions would require 5 pages of printed educational materials, at a cost of $0.13/page,[8] for a total of: 5($0.13)*6 = $3.90/person.

Based on the structure of this intervention, intervention costs we assumed to be entirely per-person with no fixed costs. The per-person cost for the 6-visit intervention combined personnel, overhead and material resource costs: $315.17+$664.32+$3.90 = $983.39.

Base case costs for each intervention are summarized in Supplemental Table 4. In combination strategies, monthly fixed costs were additive and one-time per-person costs were additive. For example, someone who engaged in both the initiation and persistence strategy incurred intervention costs of $172.34+$679.25=$851.59 while the fixed costs for the combination initiation & persistence intervention were $1,659.54+$1,325.24=$2,984.78 per month regardless of the number of people enrolled in the interventions.

**Supplemental Table 4.** Base Case Intervention Costs

| Category | Initiation:<br>Health Mindr | Persistence: PrEP@Home and ePrEP | Adherence:<br>Life-Steps PrEP |
|---|---|---|---|
| **Personnel** | Fixed: $1,075.46 | Fixed: $741.16<br>Per Person: $243.49 | Per Person: $315.17 |
| **Overhead** | $0 | $0 | Per Person: $664.32 |
| **Direct Medical** | Per Person: $29.34 | Per Person: $435.72 | $0 |
| **Technology/material resources** | Fixed: $584.08<br>Per Person: $84.32 | Fixed: $584.08 | Per Person: $3.90 |
| **Monthly Fixed Total** | **$1,659.54** | **$1,325.24** | **$0** |
| **One-Time Per Person** | **$172.34** | **$679.25** | **$983.39** |

##### 11.7 PrEP and Other Healthcare Cost Estimates

The cost of traditional PrEP consisted of both medication costs and the cost of required medical care. Monthly PrEP drug costs (30 pills, $1,381) were obtained from US Department of Veterans Affairs Federal Supply Schedule for pharmaceuticals.[11]Clinic visits and lab costs required for traditional PrEP (i.e. PrEP not supported through the persistence intervention) were estimated from Centers for Medicare and Medicaid Services (CMS) Fee Schedules (Supplemental Table 5).[9] Guidelines recommend screening for syphilis, gonorrhea and chlamydia for MSM on PrEP.[10] For gonorrhea and chlamydia, guidelines recommend nucleic acid amplification tests (NAAT) with 3-site testing (urine, rectal, pharyngeal). Testing is done at PrEP initiation and every 6 months at follow-up clinic visits for people using PrEP.

Annual costs of medical care for HIV-positive MSM were derived from two sources to estimate costs by HIV status, treatment status, and age (Supplemental Table 6). Enns *et al.* (2019) reported quarterly costs for MSM in the South by CD4 count.[12] Schackman *et al.* (2015) reported costs by age and CD4 count.[13] Healthcare costs for HIV-negative MSM by age were obtained from median expenditure data published by the Agency for Healthcare Research and Quality (Supplemental Table 7).[14]

**Supplemental Table 5.** Clinic and Laboratory Costs

| Service | Cost | Code | Description | Source |
|---|---|---|---|---|
| Clinic visit at PrEP initiation | $187.89 | CPT 99203 | 30-minute evaluation and management visit for new patient in hospital outpatient facility | CMS Fee Schedule; NASTAD Billing Guide |
| Clinic visit for PrEP follow-up | $163.01 | CPT 99213 | 15-minute evaluation and management visit for established patient in hospital outpatient facility | |
| HIV+ Post-Test Counseling | $191.14 | CPT 99214 | 25-minute evaluation and management visit for established patient in hospital outpatient facility | |
| Creatinine Clearance Test | $9.46 | HCPCS 82575 | Creatinine clearance test | CMS Clinical Laboratory Fee Schedule 2020 |
| Hepatitis B Test | $10.33 | HCPCS 87340 | Hepatitis B surface antigen | |
| Rapid HIV Test | $13.71 | HCPCS 86703 | HIV-1/HIV-2 1 result antibody | |
| Confirmatory HIV Test | $22.41 | HCPCS 86702, 86701 | HIV-2 antibody and HIV-1 antibody | |
| Syphilis Test | $4.27 | HCPCS 86592 | Syphilis test non-trep qual | |
| Chlamydia Test (NAAT) | $35.09 | HCPCS 87491 | Chylmd. Trach. DNA amp probe | |
| Gonorrhea Test (NAAT) | $35.09 | HCPCS 87591 | N.gonorrhoeae DNA amp prob | |

CMS: Centers for Medicare and Medicaid Services; NASTAD: National Alliance of State and Territorial AIDS Directors; CPT: Current Procedural Terminology; HCPCS: Healthcare Common Procedure Coding System; NAAT: nucleic acid amplification tests.

**Supplemental Table 6.** Annual Costs of Medical Care for HIV-Positive Individuals*

| HIV Stage and Treatment Status | Age (years) | | | |
|---|---|---|---|---|
| | 18-29 | 30-39 | 40-49 | 50+ |
| **HIV+, Not on ART** | $5,066.04 | $5,836.15 | $6,426.64 | $6,925.30 |
| **HIV+, On ART** | $40,365.96 | $46,502.22 | $51,207.15 | $55,180.51 |
| **AIDS, Not on ART** | $11,790.88 | $13,583.03 | $14,957.43 | $16,118.02 |
| **AIDS, On ART** | $43,546.14 | $50,164.94 | $55,240.86 | $59,527.15 |

*Includes HIV-related and non-HIV medical costs. Reported in 2020 USD.

**Supplemental Table 7.** Annual General Medical Costs for HIV-Negative Men

| | Age (years) | | |
|---|---|---|---|
| | 18-44 | 45-64 | 65+ |
| **Annual Cost** | $2,304.45 | $7,063.02 | $12,761.69 |

Source: Agency for Healthcare Research and Quality. Median expenditure per person with expense (standard errors) by age groups and sex, United States, 2017. Medical Expenditure Panel Survey. Generated interactively: Sun Mar 15 2020. https://meps.ahrq.gov/mepstrends/hc_use/

#### 11.8 Impact Inventory

The impact inventory outlines what effects and costs were included in the cost-effectiveness analysis, by sector. The analysis was conducted from a healthcare sector perspective and therefore did not include costs or effects in sectors other than the formal healthcare sector. However, the impact inventory briefly discusses the expected impact of the interventions in these other sectors.

| Sector | Type of Impact | Included in Analysis | | Notes of Evidence |
|---|---|---|---|---|
| | | Health Sector | Payer | |
| **HEALTH** | ***Formal Healthcare Sector*** | | | |
| | Longevity effects | Yes | Yes | |
| | Health-related quality of life (HRQoL) effects | Yes | Yes | |
| | Infection transmission | Yes | Yes | Transmission tracked for 20-yrs |
| | Adverse events | No | No | Assumed to be part of HRQoL |
| | Intervention costs | Yes | Yes | Payer is assumed to be a funder, like a health department |
| | PrEP drug costs | Yes | No | |
| | Testing and clinic visit costs | Yes | No | |
| | Future related medical costs | Yes | No | |
| | Future unrelated medical costs | Yes | No | |
| | ***Informal Healthcare Sector***<br>*Not considered because did not conduct societal perspective analysis* | | | |
| | Patient time costs | **Persistence** intervention expected to have lower patient time costs than status quo because of avoided transportation time and clinic wait time with home-based PrEP.<br>**Adherence** intervention would incur a patient time cost for the six in-clinic adherence counseling session as well as transport to the clinic to attend.<br>**Initiation** intervention would take patient time to engage in the app and take monthly PrEP eligibility assessments but these time expenditures are expected to be negligible. | | |
| | Unpaid caregiver time | Interventions expected to have minimal impact. | | |
| | Transportation costs | **Persistence** intervention expected to have lower transportation costs than status quo because of home-based testing and PrEP visits.<br>**Adherence** expected have higher transportation costs because of additional clinic visits for intervention.<br>**Initiation** intervention expected to have minimal impact; perhaps slightly lower costs if patients do home-based testing instead of in-clinic testing | | |
| | ***Non-Healthcare Sectors***<br>*Not considered because did not conduct societal perspective analysis* | | | |
| **PRODUCTIVITY** | Labor market earnings | Expected to have minimal impact | | |
| | Cost of unpaid lost productivity due to illness or missed work | Averted HIV transmissions may reduce lost productivity since HIV as a chronic condition requires lifelong management and causes significant illness if not well-managed | | |
| | Cost of uncompensated household production | Expected to have minimal impact | | |
| **CONSUMPTION** | Future consumption unrelated to health | Interventions may result in higher future consumption to due longer life expectancy for people with averted HIV infections, although this is expected to be small since life expectancy for HIV+ and HIV- are now relatively similar if HIV is well-controlled. | | |
| **SOCIAL SERVICES** | Averted HIV transmissions may result in fewer social service needs since HIV-positive individuals often receive case management and other support services. | | | |
| **OTHER** | Interventions expected to have minimal impact on education, housing, legal, criminal justice, and environmental sectors. | | | |

#### 12 ADDITIONAL ANALYSIS RESULTS

##### 12.1 Estimating Uncertainty

The stochastic nature of the simulation model translated into high variability of model outcomes, making it difficult to estimate stable incremental differences in costs and QALYs between certain strategies. A prominent source of the stochastic variability in the model outcomes was the population size over the course of the simulation. Although all simulations started with the same initial population size, a random number of new arrivals entered the open population at each timestep, resulting in stochastic variation in population size across simulations. This stochastic process induces variability in total costs and QALYs across the simulations that is largely unrelated to the effects of the PrEP care cascade strategies being evaluated. To adjust for this stochastic variability, a simple linear regression model for each strategy was used to estimate the mean discounted costs and QALYs for each simulation conditional on the mean number of individuals alive during the simulated time horizon for that simulation. Using this regression model, costs and QALYs for each simulation were standardized to reflect a single average population size.

Outcome point estimates reported in Table 2 of the main text reflect mean population-size adjusted discounted costs and QALYs averaged across 10,000 independent simulations of the model for each strategy using base case parameter inputs. To capture how model stochasticity translates into uncertainty in our cost-effectiveness results, bootstrapping was used to estimate the sampling distribution of ICER values along the cost-effective frontier. The bootstrapped ICER distribution was generated by drawing 1,000 samples of size 10,000 with replacement from the population-size adjusted cost and QALY outcome distributions for strategies along the efficient frontier (i.e., strategies “-A-”, “IA-”, and “IAP”). For each set of the 1,000 samples, an ICER was calculated as the difference in sample mean costs divided by the difference in sample mean QALYs between the relevant strategies. The 2.5$^{th}$ and 97.5$^{th}$ quantiles of the resulting distribution of 1,000 ICER values define the confidence interval for the ICER of interest. For the “-A-” strategy, characterizing ICER uncertainty was challenging because of the small benefit it conferred. Though the mean incremental QALYs gained under the “-A-” strategy (relative to the status quo) was 141 QALYs, there was a substantial proportion (16.2%) of the bootstrapped samples for which the “-A-” strategy had fewer QALYs than under the status quo (Supplemental Figure 1). Though there is no mechanism in the simulation model by which the adherence intervention could be harmful to PrEP users, this demonstrates that the modest benefits of the adherence intervention can be overwhelmed by the stochastic variation in the simulated outcomes.

**Supplemental Figure 1.** Bootstrap Means for Incremental Costs and QALYs, Comparing Status Quo and Adherence Strategies.

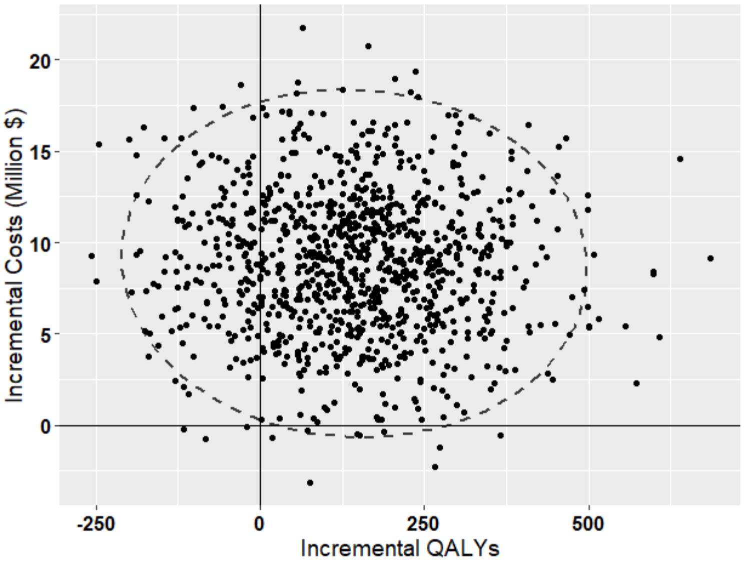

##### 12.2 Intervention Coverage Sensitivity Analysis

We varied the proportion of participants engaging in the interventions to explore how assumptions regarding intervention capacities impact cost-effectiveness results (Supplemental Table 8). Coverage levels were mainly chosen to represent a range of potential scale-up levels, based loosely on assumed acceptability or maximum capacity of interventions. For example, monthly per-person and fixed costs for interventions were kept at base case levels (i.e., we did not assume additional costs for reaching higher proportions of participants when considering higher coverage levels).

**Supplemental Table 8.** Intervention Coverage Scenario Assumptions

| Intervention | Low | Base Case | High |
|---|---|---|---|
| **Initiation**, % of HIV- and undiagnosed HIV+ MSM using intervention app | 12.5% | 22.5% | 30.5% |
| **Adherence**, % of MSM starting PrEP engaged in intervention | 15% | 30% | 60% |
| **Persistence**, % of MSM starting PrEP engaged in intervention | 20% | 40% | 80% |

Strategy incremental costs and QALYs relative to the status quo ($45,737 million and 3,125,390 QALYs, respectively) are presented for the difference coverage levels in Supplemental Table 9. In the low coverage scenario, effects of the adherence intervention were largely undetectable in our simulations. Effect sizes were also relatively small for the other strategies, making results subject to a fair amount of model stochasticity. The initiation & adherence combination strategy was still found to be cost-effective in this low coverage scenario, but with a high degree of variability. In the high coverage scenario, cost-effectiveness results were similar to those found in the base case, with A, IA, and IAP on the efficient frontier and IA considered to be cost-effective with an ICER of $90,338/QALY gained. The similarity in ICERs for strategies under base case and high coverage assumptions indicates that for these coverage levels, costs and benefits are scaling approximately linearly with the number of individuals reached by the interventions. As expected, high coverage scenarios did achieve more substantial gains in PrEP coverage, infections averted, and QALYs gained than under base case coverage assumptions.

| **Supplemental Table 9.** Results of Scenario Analyses on Intervention Coverage | | | | | | | | |
|---|---|---|---|---|---|---|---|---|
| **Intervention** | **PrEP Coverage (%)** | **Infections Averted (%)** | **Difference from Status Quo** | | **Incremental Differences** | | **ICER ($/QALY gained)** | **ICER Bootstrap 95% CI** |
| | | | **Cost (millions)** | **QALYs** | **Cost (millions)** | **QALYs** | | |
| **Low Intervention Coverage*** | | | | | | | | |
| Status Quo | 15.1 | -- | -- | -- | -- | -- | -- | |
| Initiation & Adherence (IA) | 16.9 | 1.22 | 231.5 | 2,468 | 231.5 | 2,468 | 93,774 | (85,647, 110,255) |
| Initiation & Persistence (IP) | 18.1 | 1.55 | 306.2 | 2,875 | 74.7 | 407 | 183,652 | (108,313, 632,637) |
| Adherence & Persistence (AP) | 16.2 | 0.45 | 79.4 | 615 | -- | -- | ED | |
| Initiation (I) | 16.9 | 1.14 | 219.9 | 2,257 | -- | -- | ED | |
| Adherence (A) | 15.1 | 0.08 | 5.0 | 0* | -- | -- | D | |
| Persistence (P) | 16.2 | 0.30 | 8.1 | 354 | -- | -- | D | |
| Combination (IAP) | 18.1 | 1.66 | 310.4 | 2,865 | -- | -- | D | |
| **High Intervention Coverage Scenario** | | | | | | | | |
| Status Quo | 15.1 | -- | -- | -- | -- | -- | -- | |
| Adherence (A) | 15.1 | 0.41 | 19.1 | 612.1 | 19.1 | 612.1 | 31,177 | (17,433, 67,579) |
| Initiation & Adherence (IA) | 22.7 | 5.10 | 815.7 | 9,430.6 | 796.6 | 8,818.5 | 90,338 | (87,537, 93,353) |
| Combination (IAP) | 28.2 | 7.16 | 1,185.5 | 11,625.3 | 369.8 | 2,194.8 | 168,484 | (149,232, 195,339) |
| Persistence (P) | 19.3 | 1.54 | 294.7 | 2,003.8 | -- | -- | ED | |
| Adherence & Persistence (AP) | 19.3 | 1.99 | 305.9 | 2,521.1 | -- | -- | ED | |
| Initiation (I) | 22.6 | 4.40 | 784.8 | 8,627.3 | -- | -- | ED | |
| Initiation & Persistence (IP) | 28.1 | 6.47 | 1,162.6 | 10,967.5 | -- | -- | ED | |

QALY: Quality-adjusted life year; ICER: incremental cost-effectiveness ratio, reported as cost/QALY gained; CI: confidence interval; ED: extended dominance, meaning the linear combination of two other strategies is more efficient than the labeled strategy. D: strong dominance, meaning the strategy produces fewer QALYs at a higher cost than another strategy. Costs are reported in 2020 USD.

* Estimates for low coverage scenarios, particularly for strategies including adherence, were unstable due to low effect size and model stochasticity.

##### 12.3 Payer Perspective Analysis

In the payer perspective analysis, we assume a federal or state agency payer who would be funding interventions to address PrEP barriers and/or support PrEP use, without directly paying for the cost of PrEP medication or medical care. Therefore, in this analysis, only intervention costs are included. The payer perspective was only assessed in the base case and high coverage scenarios due to the instability of results in the low coverage scenario (as described above). Under a payer perspective, the cost of the status quo strategy was zero because no interventions implemented. For all strategies, QALYs remain the same as under the health sector perspective.

In the base case, initiation alone (ICER $19,555/QALY gained), initiation & persistence combination ($31,397/QALY gained), and the triple combination (ICER $70,684) were non-dominated (Supplemental Table 10). This differs from the healthcare perspective, where strategies involving the adherence intervention were typically non-dominated. This is because the adherence intervention, while only conferring a small benefit, was inexpensive relative to the other interventions because it did not increase the number of users on PrEP and incur the associated increase in PrEP costs. However, when PrEP costs are not considered, the difference in cost between the adherence intervention and the initiation and persistence interventions is much smaller. This makes the initiation and persistence interventions much more efficient, achieving much higher numbers of HIV infections averted and associated QALY gains than the adherence intervention. A similar pattern is seen in the high intervention coverage scenario.

| **Supplemental Table 10.** Cost-Effectiveness Results Using a Payer Perspective | | | | |
|---|---|---|---|---|
| **Intervention & Coverage Scenario** | **Cost (2020 USD, millions)** | **Incremental Differences** | | **ICER ($/QALY gained)** |
| | | **Cost (millions)** | **QALYs** | |
| **Base Case Intervention Coverage** | | | | |
| Status Quo | 0 | -- | -- | -- |
| Initiation (I) | 112.7 | 112.7 | 5,764 | 19,555 |
| Initiation & Persistence (IP) | 146.9 | 34.2 | 1,089 | 31,397 |
| Combination (IAP) | 184.4 | 37.4 | 530 | 70,684 |
| Persistence (P) | 27.6 | -- | -- | ED |
| Adherence & Persistence (AP) | 56.2 | -- | -- | ED |
| Adherence (A) | 29.1 | -- | -- | D |
| Initiation & Adherence (IA) | 151.2 | -- | -- | D |
| **High Intervention Coverage Scenario** | | | | |
| Status Quo | 0 | -- | -- | -- |
| Initiation (I) | 150.7 | 150.7 | 8,627 | 17,470 |
| Initiation & Persistence (IP) | 223.2 | 72.5 | 2,340 | 30,971 |
| Combination (IAP) | 305.7 | 82.5 | 658 | 125,391 |
| Persistence (P) | 52.8 | -- | -- | ED |
| Adherence & Persistence (AP) | 108.8 | -- | -- | ED |
| Adherence (A) | 58.3 | -- | -- | D |
| Initiation & Adherence (IA) | 238.5 | -- | -- | D |

QALY: Quality-adjusted life year; ICER: incremental cost-effectiveness ratio, reported as cost/QALY gained; ED: extended dominance, meaning the linear combination of two other strategies is more efficient than the labeled strategy. D: strong dominance, meaning the strategy produces fewer QALYs at a higher cost than another strategy.

#### 13 REFERENCES: SECTIONS 11-12
